# Six-month Neurological and Psychiatric Outcomes in 236,379 Survivors of COVID-19

**DOI:** 10.1101/2021.01.16.21249950

**Authors:** M. Taquet, J.R. Geddes, M. Husain, S. Luciano, P.J. Harrison

## Abstract

**Background:** Neurological and psychiatric sequelae of COVID-19 have been reported, but there are limited data on incidence rates and relative risks.

**Methods:** Using retrospective cohort studies and time-to-event analysis, we estimated the incidence of ICD-10 diagnoses in the 6 months after a confirmed diagnosis of COVID-19: intracranial haemorrhage; ischaemic stroke; Parkinsonism; Guillain-Barré syndrome; nerve/nerve root/plexus disorders; myoneural/muscle disease; encephalitis; dementia; mood, anxiety, and psychotic disorders; substance misuse; and insomnia. Data were obtained from the TriNetX electronic health records network (over 81 million patients). We compared incidences with those in propensity score-matched cohorts of patients with influenza or other respiratory infections using a Cox model. We investigated the effect on incidence estimates of COVID-19 severity, as proxied by hospitalization and encephalopathy (including delirium and related disorders).

**Findings:** 236,379 patients survived a confirmed diagnosis of COVID-19. Among them, the estimated incidence of neurological or psychiatric sequelae at 6 months was 33.6%, with 12.8% receiving their first such diagnosis. Most diagnostic categories were commoner after COVID-19 than after influenza or other respiratory infections (hazard ratios from 1.21 to 5.28), including stroke, intracranial haemorrhage, dementia, and psychotic disorders. Findings were equivocal for Parkinsonism and Guillain-Barré syndrome. Amongst COVID-19 cases, incidences and hazard ratios for most disorders were higher in patients who had been hospitalized, and markedly so in those who had experienced encephalopathy. Results were robust to sensitivity analyses, including comparisons against an additional four index health events.

**Interpretation:** The study provides evidence for substantial neurological and psychiatric morbidity following COVID-19 infection. Risks were greatest in, but not limited to, those who had severe COVID-19. The information can help in service planning and identification of research priorities.

**Funding:** National Institute for Health Research (NIHR) Oxford Health Biomedical Research Centre.

---

Since the COVID-19 pandemic began, there has been concern that survivors might be at an increased risk of neurological disorders. This concern, initially based on findings from other coronaviruses,^1^ was followed rapidly by case series,^2-4^ emerging evidence of COVID-19 CNS involvement,^5-7^ and the identification of mechanisms by which this could occur.^8-11^ There have been similar concerns regarding psychiatric sequelae of COVID-19,^12,13^ with evidence showing that survivors are indeed at increased risk of mood and anxiety disorders, as well as dementia, in the three months after infection.^14^ However, large-scale, robust, and longer-term data are needed if the consequences of the COVID-19 pandemic on brain health are to be identified and quantified. Such information is required both to plan services and to identify research priorities. We used an electronic health records network to investigate the incidence of neurological and psychiatric diagnoses in survivors in the six months after documented clinical COVID-19 infection, and the hazard ratios compared to other health conditions. We explored whether hospitalization and encephalopathy during acute COVID-19 illness affect these risks. The trajectory of hazard ratios across the six-month period was also investigated.

## Methods

### Data and study design

The TriNetX Analytics Network (www.trinetx.com) is a federated network recording anonymized data from electronic health records in 62 healthcare organizations, totalling 81 million patients. The network and its functionalities have been described elsewhere.^12^ Available data include demographics, diagnoses (using ICD-10 codes), medications, procedures, and measurements (e.g. blood pressure). The healthcare organizations are a mixture of hospitals, primary care, and specialist providers and contribute data from uninsured as well as insured patients. Using the TriNetX user interface, cohorts can be created based on inclusion and exclusion criteria, matched for confounding variables using a built-in propensity score matching algorithm, and compared for outcomes of interest over specified time periods. TriNetX has a waiver from the Western Institutional Review Board since only aggregated counts and summaries of de-identified information are used. Further details are provided in the Appendix (p. 1).

### Cohorts

The primary cohort was defined as all patients who had a confirmed diagnosis of COVID-19 (ICD-10 code U07.1). We also produced two matched control cohorts: patients diagnosed with influenza (ICD-10 codes J09-J11) and patients diagnosed with any respiratory tract infection (ICD-10 codes J00–J06, J09–J18, or J20–J22). We excluded patients with a diagnosis of COVID-19 or a positive test for COVID-19 from the control cohorts. We refer to the diagnosis of COVID-19 (in the primary cohort), influenza, or other respiratory tract infections (in the control cohorts) as index events. All cohorts included all patients over the age of ten who had the index event on or after January 20, 2020 (the date of the first recorded COVID-19 case in the USA) and who were still alive at the time of the analysis (December 13, 2020). Further details on cohorts are provided in the Appendix (p. 1).

### Covariates

A set of established and suspected risk factors for COVID-19 and for more severe COVID-19 illness was used:^15,16^ age, sex, race, ethnicity, obesity, hypertension, diabetes, chronic kidney disease, asthma, chronic lower respiratory diseases, nicotine dependence, substance misuse, ischaemic heart disease and other forms of heart disease, socioeconomic deprivation, cancer (and haematological cancer in particular), chronic liver disease, stroke, dementia, organ transplant, rheumatoid arthritis, lupus, psoriasis, and other immunosuppression. To capture these risk factors in patients’ health records, 55 variables were used. More details are provided in the Appendix (pp. 2-3). Cohorts were matched for all these variables, as described below.

### Outcomes

We investigated neurological and psychiatric sequelae of COVID-19 in terms of 14 outcomes occurring in the period from 1 to 180 days after the index event: intracranial haemorrhage (ICD-10 codes I60–I62); ischaemic stroke (I63); Parkinson’s disease and Parkinsonism (G20– G21); Guillain-Barré syndrome (G61.0); nerve, nerve root, and plexus disorders (G50–G59); myoneural junction and muscle disease (G70–G73); encephalitis (G04, G05, A86, or A85.8); dementia (F01–F03, G30, G31.0, or G31.83); psychotic, mood, or anxiety disorder (F20–F48) as well as each category separately; substance use disorder (F10–F19), and insomnia (F51.0 or G47.0).

For outcomes that are chronic illnesses (e.g. dementia, Parkinson’s disease), patients who had the diagnosis before the index health event were excluded. For outcomes that tend to recur or relapse (e.g. ischaemic strokes, psychiatric diagnoses), we estimated separately the incidence of *first* diagnoses (i.e. excluding those who had a diagnosis before the index event) and the incidence of *any* diagnosis (i.e. including patients who had a diagnosis at some point before the index event). For other outcomes (e.g. Guillain-Barré syndrome), the incidence of *any* diagnosis was estimated. More details are provided in the Appendix (p. 3).

Finally, to assess the overall risk of neurological and psychiatric outcomes after COVID-19, we estimated the incidence of any of the 14 outcomes, and the incidence of a first diagnosis of any of the outcomes. Note that this is less than the sum of the incidences of each outcome since some patients have more than one diagnosis.

### Secondary analyses

We investigated whether the neurological and psychiatric sequelae of COVID-19 is affected by the severity of the illness in three ways. The incidence of outcomes was estimated separately among patients who: (a) had required hospitalization, within a time window from 4 days before their COVID-19 diagnosis (taken to be the time it might take between clinical presentation and confirmation) to 2 weeks afterwards; (b) had not required hospitalization during that window, and (c) patients who were diagnosed with delirium or other forms of altered mental status during that window; here, we use the term ‘encephalopathy’ to describe this group of patients (see Appendix p. 3 for list of ICD-10 codes).^17,18^

Differences in outcome incidence between these subgroups might reflect differences in their baseline characteristics. For each outcome, we therefore estimated the HR between patients requiring hospitalization and a matched cohort of patients not requiring hospitalization, and between patients with encephalopathy and a matched cohort of patients without encephalopathy. Finally, HRs were calculated for patients who had not required hospitalization for COVID-19, influenza, or other respiratory infections.

To provide benchmarks for the incidence of neurological and psychiatric sequelae, HRs for COVID-19 were compared to four additional matched cohorts of patients diagnosed with health events selected to represent a range of acute presentations during the same time period. The health events were: (i) skin infection, (ii) urolithiasis, (iii) fracture of a large bone, and (iv) pulmonary embolism. For details, see Appendix (p. 4).

To test whether differences in sequelae between cohorts could be accounted for by differences in follow up, we counted the average number of health visits that each cohort had during the follow-up period.

### Statistical analyses

Propensity score matching^19^ was used to create cohorts with matched baseline characteristics, and carried out within the TriNetX network. Propensity score 1:1 matching used a greedy nearest neighbour matching approach with a caliper distance of 0.1 pooled standard deviations of the logit of the propensity score. Any characteristic with a standardized mean difference (SMD) between cohorts lower than 0.1 is considered well matched.^20^ The incidence of each outcome was estimated using the Kaplan-Meier estimator. Comparisons between cohorts were made using a log-rank test. Hazard ratios (HR) were calculated using a proportional hazard model wherein the cohort to which the patient belonged was used as the independent variable. The proportional hazard assumption was tested using the generalized Schoenfeld approach. When the assumption was violated, the time-varying HR was assessed using natural cubic splines fitted to the log cumulative hazard.^21^ See Appendix (p. 4) for further details.

Statistical analyses were conducted in R version 3.4.3 except for the log-rank tests which were performed within TriNetX. Statistical significance was set at two-sided p-values < 0⋅05.

## Results

Using electronic health records, the incidence of neurological and psychiatric diagnoses was measured in the six months after COVID-19 infection, and compared to propensity-score-matched cohorts of patients with influenza or other respiratory tract infections. We explored how incidences and HRs differed according to hospitalization and encephalopathy during the acute illness, and how HRs changed over the six months. Key findings are summarised here, with additional details, results, and sensitivity analyses, shown in the Appendix. Adequate propensity-score matching (SMD≤0.1) was achieved for all comparisons and baseline characteristics.

Table 1 (and Table S1 in the Appendix) summarises the main demographic features and comorbidities of the COVID-19 cohort (n=236,379), as well as the subgroups who were not-hospitalized (n=190,077), hospitalized (n=46,302), or who had a diagnosis of encephalopathy (n=6,229). Table 1 also presents the estimated diagnostic incidence of the major neurological and psychiatric outcomes over the following six months. All diagnoses are commoner in those who had been hospitalized, and markedly so in those who were encephalopathic during the illness. Results according to gender, race and age are shown in the Appendix (Table S2).

**Table 1:** Baseline characteristics and major outcomes for the whole COVID-19 cohort, the non-hospitalized and hospitalized sub-groups, and those with encephalopathy during the illness. Only characteristics with a prevalence higher than 5% in the whole population are displayed. For additional baseline characteristics and outcomes see Table S1.

|  | All patients | Non-hospitalized patients | Hospitalized patients | Patients with encephalopathy |
| --- | --- | --- | --- | --- |
| **COHORT SIZE** | 236379 (100.0) | 190077 (100.0) | 46302 (100.0) | 6229 (100.0) |
| **DEMOGRAPHICS** |  |  |  |  |
| Age, mean (SD), y | 46 (19.7) | 43.3 (19.0) | 57 (18.7) | 66.7 (17.0) |
| Sex, n (%) females | 131460 (55.6) | 107730 (56.7) | 23730 (51.2) | 2909 (46.7) |
| Race, n (%) |  |  |  |  |
| White | 135143 (57.2) | 109635 (57.7) | 25508 (55.1) | 3331 (53.5) |
| Black or African American | 44459 (18.8) | 33868 (17.8) | 10591 (22.9) | 1552 (24.9) |
| Unknown | 48085 (20.3) | 39841 (21.0) | 8244 (17.8) | 1071 (17.2) |
| Ethnicity, n (%) |  |  |  |  |
| Hispanic or Latino | 37772 (16.0) | 29155 (15.3) | 8617 (18.6) | 895 (14.4) |
| Not Hispanic of Latino | 134075 (56.7) | 106844 (56.2) | 27231 (58.8) | 3873 (62.2) |
| Unknown | 64532 (27.3) | 54078 (28.5) | 10454 (22.6) | 1461 (23.5) |
| **COMORBIDITIES, n (%)** |  |  |  |  |
| Overweight and obesity | 42871 (18.1) | 30198 (15.9) | 12673 (27.4) | 1838 (29.5) |
| Hypertensive disease | 71014 (30.0) | 47516 (25.0) | 23498 (50.7) | 4591 (73.7) |
| Type 2 diabetes mellitus | 36696 (15.5) | 22518 (11.8) | 14178 (30.6) | 2890 (46.4) |
| Asthma | 25104 (10.6) | 19834 (10.4) | 5270 (11.4) | 755 (12.1) |
| Nicotine dependence | 17105 (7.2) | 12639 (6.6) | 4466 (9.6) | 803 (12.9) |
| Substance misuse | 24870 (10.5) | 18173 (9.6) | 6697 (14.5) | 1316 (21.1) |
| Ischaemic heart diseases | 21082 (8.9) | 11815 (6.2) | 9267 (20.0) | 2200 (35.3) |
| Other forms of heart disease | 42431 (18.0) | 26066 (13.7) | 16365 (35.3) | 3694 (59.3) |
| Chronic kidney disease (CKD) | 15908 (6.7) | 8345 (4.4) | 7563 (16.3) | 1892 (30.4) |
| Neoplasms | 45255 (19.1) | 34362 (18.1) | 10893 (23.5) | 1793 (28.8) |
| **OUTCOMES, % at 6 months (95% CI)** |  |  |  |  |
| Intracranial haemorrhage (any) | 0.56 (0.50-0.63) | 0.31 (0.25-0.39) | 1.31 (1.14-1.52) | 3.61 (2.97-4.39) |
| Intracranial haemorrhage (first) | 0.28 (0.23-0.33) | 0.14 (0.10-0.20) | 0.63 (0.50-0.80) | 1.19 (0.82-1.70) |
| Ischaemic stroke (any) | 2.10 (1.97-2.23) | 1.33 (1.22-1.46) | 4.38 (4.05-4.74) | 9.35 (8.23-10.62) |
| Ischaemic stroke (first) | 0.76 (0.68-0.85) | 0.43 (0.36-0.52) | 1.60 (1.37-1.86) | 3.28 (2.51-4.27) |
| Parkinsonism | 0.11 (0.079-0.14) | 0.074 (0.046-0.12) | 0.20 (0.15-0.28) | 0.46 (0.28-0.78) |
| Guillain-Barre syndrome | 0.081 (0.062-0.11) | 0.047 (0.03-0.072) | 0.22 (0.15-0.32) | 0.48 (0.20-1.14) |
| Nerve/nerve root/plexus disorders | 2.85 (2.69-3.03) | 2.69 (2.51-2.89) | 3.35 (3.02-3.72) | 4.69 (3.81-5.77) |
| Myoneural junction/muscle disease | 0.45 (0.40-0.52) | 0.16 (0.12-0.20) | 1.24 (1.05-1.46) | 3.27 (2.54-4.21) |
| Encephalitis | 0.10 (0.076-0.13) | 0.05 (0.032-0.076) | 0.24 (0.17-0.33) | 0.64 (0.39-1.07) |
| Dementia | 0.67 (0.59-0.75) | 0.35 (0.29-0.43) | 1.46 (1.26-1.71) | 4.72 (3.80-5.85) |
| Mood/Anxiety/Psychotic disorder (any) | 23.98 (23.58-24.38) | 23.59 (23.12-24.07) | 24.50 (23.76-25.26) | 36.25 (34.16-38.43) |
| Mood/Anxiety/Psychotic disorder (first) | 8.63 (8.28-8.98) | 8.15 (7.75-8.57) | 8.85 (8.22-9.52) | 12.96 (11.13-15.07) |
| Mood disorder (any) | 13.66 (13.35-13.99) | 13.10 (12.73-13.47) | 14.69 (14.09-15.32) | 22.52 (20.71-24.47) |
| Mood disorder (first) | 4.22 (3.99-4.47) | 3.86 (3.60-4.14) | 4.49 (4.05-4.99) | 8.07 (6.56-9.90) |
| Anxiety disorder (any) | 17.39 (17.04-17.74) | 17.51 (17.09-17.93) | 16.40 (15.76-17.06) | 22.43 (20.65-24.34) |
| Anxiety disorder (first) | 7.11 (6.82-7.41) | 6.81 (6.47-7.16) | 6.91 (6.38-7.47) | 9.24 (7.70-11.07) |
| Psychotic disorder (any) | 1.40 (1.30-1.51) | 0.93 (0.83-1.04) | 2.89 (2.62-3.18) | 7.00 (6.01-8.14) |
| Psychotic disorder (first) | 0.42 (0.36-0.49) | 0.25 (0.19-0.33) | 0.89 (0.72-1.09) | 2.12 (1.53-2.94) |
| Substance misuse (any) | 6.58 (6.36-6.80) | 5.87 (5.63-6.13) | 8.56 (8.10-9.04) | 11.85 (10.55-13.31) |
| Substance misuse (first) | 1.92 (1.77-2.07) | 1.74 (1.58-1.91) | 2.09 (1.82-2.40) | 2.58 (1.91-3.47) |
| Insomnia (any) | 5.42 (5.20-5.64) | 5.16 (4.91-5.42) | 5.95 (5.53-6.39) | 9.82 (8.57-11.24) |
| Insomnia (first) | 2.53 (2.37-2.71) | 2.23 (2.05-2.43) | 3.14 (2.81-3.51) | 5.05 (4.10-6.20) |
| Any | 33.62 (33.17-34.07) | 31.74 (31.22-32.27) | 38.73 (37.87-39.60) | 62.34 (60.14-64.55) |
| Any first | 12.84 (12.36-13.33) | 11.51 (10.98-12.07) | 15.29 (14.32-16.33) | 31.13 (27.29-35.36) |

Table 2 summarises the HRs for COVID-19 compared to the matched cohorts with influenza (n=105,579; matched baseline characteristics in Table S3) and with other respiratory infections (n=236,038; matched baseline characteristics in Table S4); HRs for diagnostic subcategories are given in Table S5). Kaplan-Meier curves for COVID-19 compared with the other respiratory tract infections cohort are illustrated in Figure 1 (and Figures S1–S3). HRs are statistically significantly higher than 1 for all diagnoses for COVID-19 compared to influenza, except for Parkinsonism and Guillain-Barré syndrome, and are statistically significantly higher than 1 for all diagnoses relative to other respiratory tract infections. Similar results are observed when COVID-19 is compared to the four other index events (see Appendix, Table S6 and Figures S4–S7), except where an outcome has a predicted relationship to the comparator condition (e.g. stroke incidence is higher in the pulmonary embolism cohort; intracranial haemorrhage is commoner in association with fracture of a large bone).

**Table 2:** Hazard ratios for the major outcomes after COVID-19 compared to influenza and other respiratory infections (RTI). For additional details on cohort characteristics and diagnostic subcategories, see Appendix Tables S3–S5.

|  | COVID-19 vs. Influenza |  | COVID-19 vs. Other RTI |  |
| --- | --- | --- | --- | --- |
|  | (Matched cohorts n=105579) |  | (Matched cohorts n=236038) |  |
|  | HR (95% CI) | P-value | HR (95% CI) | P-value |
| **Intracranial haemorrhage (any)** | 2.44 (1.89-3.16) | <0.0001 | 1.26 (1.11-1.43) | 0.00029 |
| **Intracranial haemorrhage (first)** | 2.53 (1.68-3.79) | <0.0001 | 1.56 (1.27-1.92) | <0.0001 |
| **Ischaemic stroke (any)** | 1.62 (1.43-1.83) | <0.0001 | 1.45 (1.36-1.55) | <0.0001 |
| **Ischaemic stroke (first)** | 1.97 (1.57-2.47) | <0.0001 | 1.63 (1.44-1.85) | <0.0001 |
| **Parkinsonism** | 1.42 (0.75-2.67) | 0.19 | 1.45 (1.05-2.00) | 0.02 |
| **Guillain-Barre syndrome** | 1.21 (0.72-2.04) | 0.41 | 2.06 (1.43-2.96) | <0.0001 |
| **Nerve/nerve root/plexus disorders** | 1.64 (1.50-1.81) | <0.0001 | 1.27 (1.19-1.35) | <0.0001 |
| **Myoneural junction/muscle disease** | 5.28 (3.71-7.53) | <0.0001 | 4.52 (3.65-5.59) | <0.0001 |
| **Encephalitis** | 1.70 (1.04-2.78) | 0.028 | 1.41 (1.03-1.92) | 0.028 |
| **Dementia** | 2.33 (1.77-3.07) | <0.0001 | 1.71 (1.50-1.95) | <0.0001 |
| **Mood/Anxiety/Psychotic disorder (any)** | 1.46 (1.43-1.50) | <0.0001 | 1.20 (1.18-1.23) | <0.0001 |
| **Mood/Anxiety/Psychotic disorder (first)** | 1.81 (1.69-1.94) | <0.0001 | 1.48 (1.42-1.55) | <0.0001 |
| **Mood disorder (any)** | 1.47 (1.42-1.53) | <0.0001 | 1.23 (1.20-1.26) | <0.0001 |
| **Mood disorder (first)** | 1.79 (1.64-1.95) | <0.0001 | 1.41 (1.33-1.50) | <0.0001 |
| **Anxiety disorder (any)** | 1.45 (1.40-1.49) | <0.0001 | 1.17 (1.15-1.20) | <0.0001 |
| **Anxiety disorder (first)** | 1.78 (1.66-1.91) | <0.0001 | 1.48 (1.42-1.55) | <0.0001 |
| **Psychotic disorder (any)** | 2.03 (1.78-2.31) | <0.0001 | 1.66 (1.53-1.81) | <0.0001 |
| **Psychotic disorder (first)** | 2.16 (1.62-2.88) | <0.0001 | 1.82 (1.53-2.16) | <0.0001 |
| **Substance misuse (any)** | 1.27 (1.22-1.33) | <0.0001 | 1.09 (1.05-1.12) | <0.0001 |
| **Substance misuse (first)** | 1.22 (1.09-1.37) | 0.00064 | 0.92 (0.86-0.99) | 0.033 |
| **Insomnia (any)** | 1.48 (1.38-1.57) | <0.0001 | 1.15 (1.10-1.20) | <0.0001 |
| **Insomnia (first)** | 1.92 (1.72-2.15) | <0.0001 | 1.43 (1.34-1.54) | <0.0001 |

**Figure 1:**
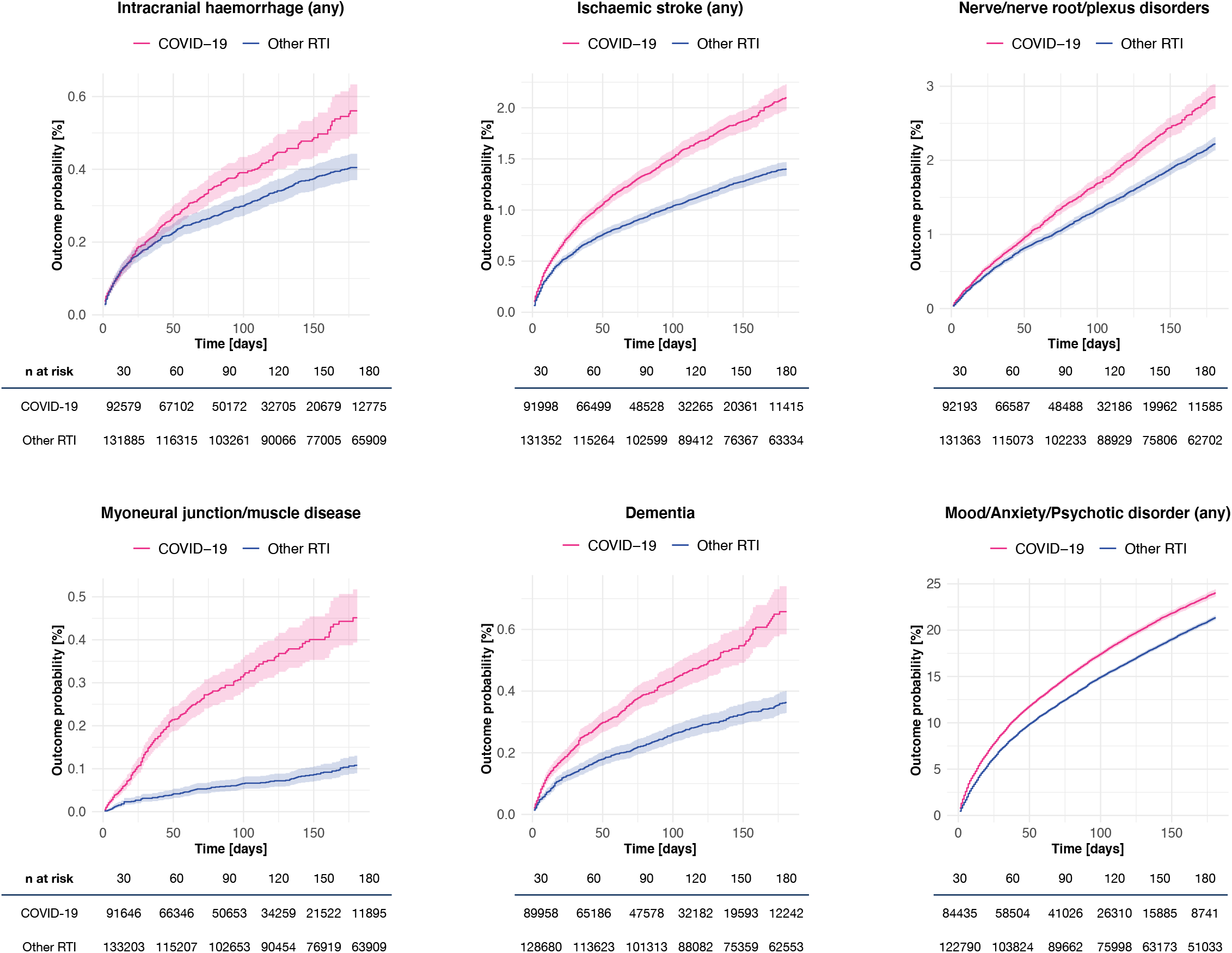
Kaplan-Meier estimates for the incidence of major outcomes after COVID-19 (pink) compared with other respiratory tract infections (blue). 95% confidence intervals are shaded. For diagnostic subcategories and additional details, see Appendix.

There were no violations of the proportional hazards assumption for most of the neurological outcomes relative to influenza or other respiratory infections over the six months (Table S7 and Figures S8–S9). The only exception is for intracranial haemorrhage or stroke when compared to other respiratory tract infections (p=0.012 and p=0.032 respectively). For the overall psychiatric disorder category (F20-F48), the HR did vary with time, declining but remaining significantly above 1, indicating that the risk is attenuated but maintained at six months. HRs for COVID-19 compared to the other four index events showed more variation with time, in part reflecting the natural history of the comparator condition (Table S8 and Figures S10–S13).

The effect of COVID-19 severity on outcomes was explored in three ways. First, by restricting analyses to matched cohorts of patients who had not required hospitalization (matched baseline characteristics in Tables S9 and S10). As shown in Table 3 (and Figure S14–S15), HRs remain significant in this subgroup. Second, by calculating the HRs for the matched hospitalized vs non-hospitalized COVID-19 cohorts (n=44,927; matched baseline characteristics in Table S11). This shows greater HRs for all outcomes in the hospitalized group, except for nerve/nerve root/plexus disorders (Table 3 and Figure 2). Third, the HRs for all diagnoses were greater for the group who had encephalopathy diagnosed during acute illness compared to a matched cohort who did not (n=6,221; matched baseline characteristics in Table S12) (Table 3 and Figure 2). HRs for the psychiatric diagnoses were less elevated in association with COVID-19 severity than were the neurological diagnoses.

**Table 3:** Hazard ratios for the major outcomes in non-hospitalized patients after COVID-19 compared to influenza or other respiratory tract infections, and for hospitalized versus non-hospitalized COVID-19 patients, and for encephalopathic versus non-encephalopathic COVID-19 patients. For details on cohort characteristics, see Appendix Tables S9–S12.

|  | COVID-19 vs. Influenza in<br>patients without hospitalization<br>(Matched cohorts n=96,803) |  | COVID-19 vs. Other RTI in<br>patients without hospitalization<br>(Matched cohorts n=183,731) |  | COVID-19 with vs. without<br>hospitalization<br>(Matched cohorts n=45,167) |  | COVID-19 with vs. without<br>encephalopathy<br>(Matched cohorts n=6,221) |  |
| --- | --- | --- | --- | --- | --- | --- | --- | --- |
|  | HR (95% CI) | P-value | HR (95% CI) | P-value | HR (95% CI) | P-value | HR (95% CI) | P-value |
| Intracranial haemorrhage (any) | 1.87 (1.25-2.78) | 0.0013 | 1.38 (1.11-1.73) | 0.0034 | 3.09 (2.43-3.94) | <0.0001 | 4.73 (3.15-7.11) | <0.0001 |
| Intracranial haemorrhage (first) | 1.66 (0.88-3.14) | 0.082 | 1.63 (1.11-2.40) | 0.01 | 3.75 (2.49-5.64) | <0.0001 | 5.00 (2.33-10.70) | <0.0001 |
| Ischaemic stroke (any) | 1.80 (1.54-2.10) | <0.0001 | 1.61 (1.45-1.78) | <0.0001 | 1.65 (1.48-1.85) | <0.0001 | 1.65 (1.38-1.97) | <0.0001 |
| Ischaemic stroke (first) | 1.71 (1.26-2.33) | 0.00031 | 1.69 (1.38-2.08) | <0.0001 | 2.82 (2.22-3.57) | <0.0001 | 3.39 (2.17-5.29) | <0.0001 |
| Parkinsonism | 2.22 (0.98-5.06) | 0.028 | 1.20 (0.73-1.96) | 0.42 | 2.63 (1.45-4.77) | 0.0016 | 1.64 (0.75-3.58) | 0.24 |
| Guillain-Barre syndrome | 0.90 (0.44-1.84) | 0.99 | 1.44 (0.85-2.45) | 0.1 | 2.94 (1.60-5.42) | 0.00094 | 2.27 (0.76-6.73) | 0.24 |
| Nerve/nerve root/plexus disorders | 1.69 (1.53-1.88) | <0.0001 | 1.23 (1.15-1.33) | <0.0001 | 0.94 (0.83-1.06) | 0.29 | 1.41 (1.07-1.87) | 0.018 |
| Myoneural junction/muscle disease | 3.46 (2.11-5.67) | <0.0001 | 2.69 (1.91-3.79) | <0.0001 | 7.76 (5.15-11.69) | <0.0001 | 5.40 (3.21-9.07) | <0.0001 |
| Encephalitis | 1.77 (0.86-3.66) | 0.095 | 2.29 (1.28-4.10) | 0.0046 | 3.26 (1.75-6.06) | 0.00017 | 9.98 (2.98-33.43) | <0.0001 |
| Dementia | 1.88 (1.27-2.77) | 0.00080 | 1.95 (1.55-2.45) | <0.0001 | 2.28 (1.80-2.88) | <0.0001 | 4.25 (2.79-6.47) | <0.0001 |
| Mood/Anxiety/Psychotic disorder (any) | 1.49 (1.45-1.54) | <0.0001 | 1.18 (1.15-1.21) | <0.0001 | 1.23 (1.18-1.28) | <0.0001 | 1.73 (1.58-1.90) | <0.0001 |
| Mood/Anxiety/Psychotic disorder (first) | 1.85 (1.72-1.99) | <0.0001 | 1.40 (1.32-1.48) | <0.0001 | 1.55 (1.40-1.71) | <0.0001 | 2.28 (1.80-2.89) | <0.0001 |
| Mood disorder (any) | 1.49 (1.43-1.55) | <0.0001 | 1.22 (1.19-1.26) | <0.0001 | 1.21 (1.15-1.28) | <0.0001 | 1.51 (1.35-1.70) | <0.0001 |
| Mood disorder (first) | 1.78 (1.61-1.96) | <0.0001 | 1.37 (1.27-1.47) | <0.0001 | 1.53 (1.33-1.75) | <0.0001 | 2.09 (1.55-2.80) | <0.0001 |
| Anxiety disorder (any) | 1.48 (1.43-1.54) | <0.0001 | 1.16 (1.13-1.19) | <0.0001 | 1.16 (1.10-1.22) | <0.0001 | 1.64 (1.45-1.84) | <0.0001 |
| Anxiety disorder (first) | 1.80 (1.67-1.94) | <0.0001 | 1.37 (1.30-1.45) | <0.0001 | 1.49 (1.34-1.65) | <0.0001 | 1.91 (1.48-2.45) | <0.0001 |
| Psychotic disorder (any) | 1.93 (1.63-2.28) | <0.0001 | 1.44 (1.27-1.62) | <0.0001 | 2.22 (1.92-2.57) | <0.0001 | 3.84 (2.90-5.10) | <0.0001 |
| Psychotic disorder (first) | 2.27 (1.56-3.30) | <0.0001 | 1.49 (1.15-1.93) | 0.0016 | 2.77 (1.99-3.85) | <0.0001 | 5.62 (2.93-10.77) | <0.0001 |
| Substance misuse (any) | 1.26 (1.19-1.33) | <0.0001 | 1.11 (1.07-1.17) | <0.0001 | 1.53 (1.42-1.64) | <0.0001 | 1.45 (1.24-1.70) | <0.0001 |
| Substance misuse (first) | 1.21 (1.05-1.38) | 0.0054 | 0.89 (0.81-0.97) | 0.013 | 1.68 (1.40-2.01) | <0.0001 | 2.03 (1.32-3.11) | 0.0015 |
| Insomnia (any) | 1.52 (1.42-1.63) | <0.0001 | 1.18 (1.12-1.24) | <0.0001 | 1.08 (0.99-1.18) | 0.088 | 1.73 (1.42-2.11) | <0.0001 |
| Insomnia (first) | 2.06 (1.82-2.33) | <0.0001 | 1.51 (1.38-1.66) | <0.0001 | 1.49 (1.28-1.74) | <0.0001 | 3.44 (2.35-5.04) | <0.0001 |

**Figure 2:**
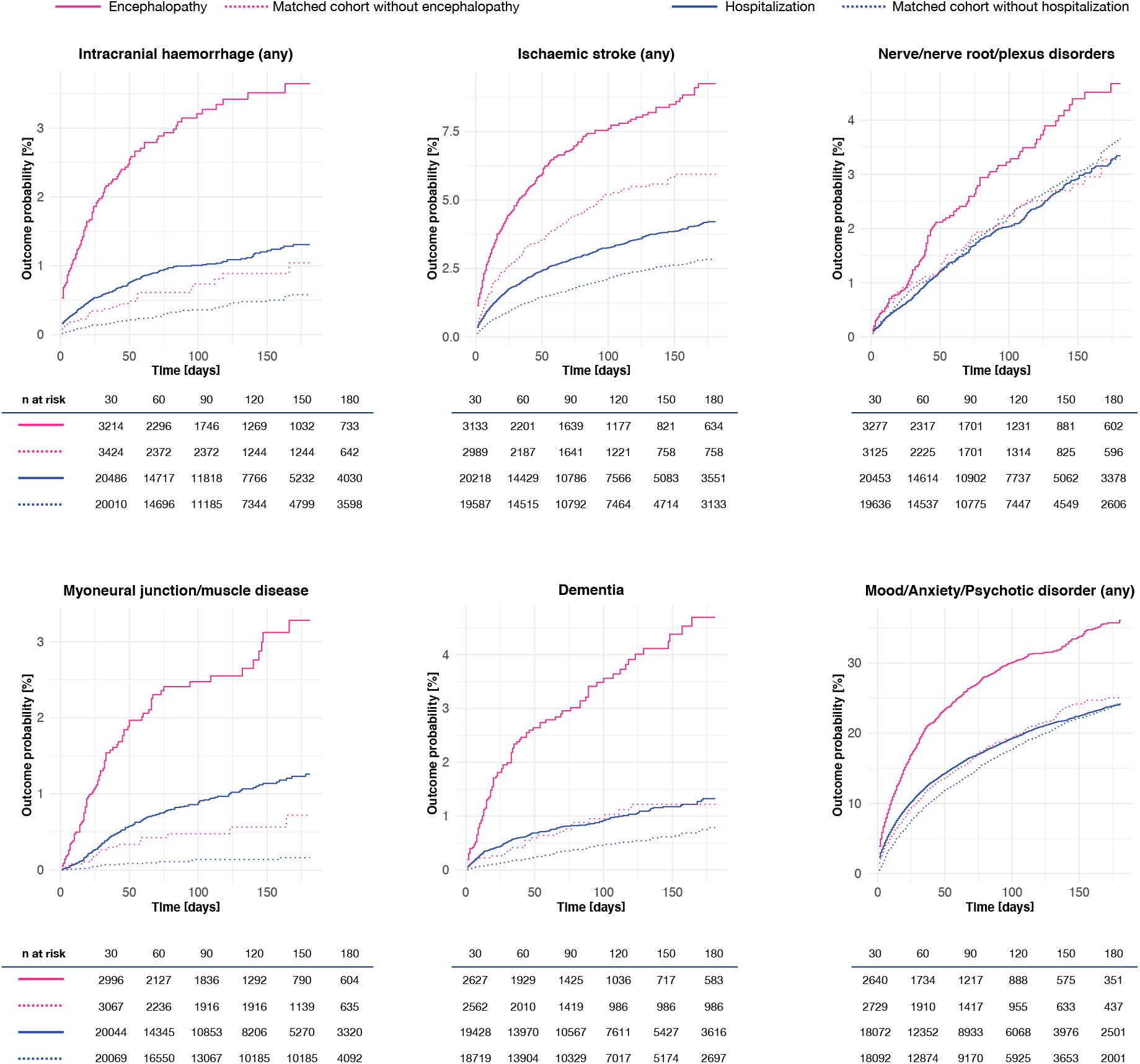
Kaplan-Meier estimates for the incidence of major outcomes after COVID-19 comparing patients requiring hospitalization (solid blue line) with matched patients not requiring hospitalization (dashed blue line), and comparing those who had encephalopathy (solid red line) with matched patients who did not have encephalopathy (dashed red line). For clarity, confidence intervals are omitted, but are shown in the Appendix, Figure S16.

We inspected for other factors that might influence the findings. (1) The results regarding hospitalization (which we had defined as occurring up to 14 days after diagnosis) could be confounded by admissions due to an early complication of COVID-19 rather than to COVID-19 itself. This was explored by excluding outcomes during this period, and findings remained, albeit with many HRs being reduced (Table S13). (2) COVID-19 survivors had fewer healthcare visits during the six-month period compared to the other cohorts (Table S14). Hence their higher incidence of many diagnoses is not simply due to having had more diagnostic opportunities.

## Discussion

A range of adverse neurological and psychiatric outcomes after COVID-19 have been predicted and reported. Data from a large electronic health records network confirm this prediction, and provide estimates of their incidence (Table 1), and the HRs compared to other matched health conditions occurring contemporaneously with the COVID-19 pandemic (Table 2 and Figure 1).

The severity of COVID-19 infection had a clear effect on subsequent neurological diagnoses (Table 3 and Figure 2). Overall, COVID-19 was associated with increased risk, but the incidences and the HRs were greater in patients who required hospitalization, and markedly so in those who developed encephalopathy, even after extensive propensity score matching for other factors (e.g. age, prior cerebrovascular disease). Potential mechanisms for this association include viral invasion of the central nervous system,^10,11^ hypercoagulable states,^22^ and neural effects of the immune response.^9^ On the other hand, it is also notable that incidence of these diagnoses was increased even in the COVID-19 cases who had not required hospitalization.

Some specific neurological diagnoses merit mention. Consistent with several other reports,^23,24^ the risk of cerebrovascular events (stroke and intracranial haemorrhage) is elevated after COVID-19, with the incidence of stroke rising to almost 1 in 10 (or 3 in 100 for a first stroke) in those who had been encephalopathic (i.e. had experienced delirium or other altered mental status; see Appendix p. 3). A similarly increased risk for stroke relative to influenza was recently reported.^25^ Whether COVID-19 is associated with Guillain-Barre syndrome has been unclear;^26^ our data indicate that the incidence is increased in hospitalized and encephalopathic patients, but not significantly in the whole cohort. There have been concerns about post-COVID-19 Parkinsonian syndromes, driven by the encephalitis lethargica epidemic that followed the 1918 influenza pandemic.^27^ Our data provide some support for this possibility although the incidence was low and not all HRs significant. It is possible that Parkinsonism is a delayed outcome, in which case a clearer signal may emerge with longer follow-up. Our previous study reported preliminary evidence for an association between COVID-19 and dementia.^14^ The present data confirm this association, with a HR of 2.33 compared to influenza, and 1.71 compared to other respiratory infections. Although the estimated incidence is modest in the whole COVID-19 cohort (0.67%), 1.46% of hospitalized cases, and 4.72% who were encephalopathic receive a first diagnosis of dementia within six months.

Our findings regarding psychiatric disorders are consistent with the 3-month outcome data we reported in a smaller number of cases using the same network,^14^ and show the HR remains elevated, although decreasing, at the six-month period. Unlike the earlier study, there is also an increased risk of psychotic disorders (likely reflecting the larger sample size and longer duration of follow-up), and substance use disorders are also more common (with the exception of the incidence of a first diagnosis of substance misuse compared to other respiratory infections). Hence, like the neurological outcomes, the psychiatric sequelae of COVID-19 appear widespread, and persist to and probably beyond six months. Compared to the neurological disorders, HRs for the common psychiatric disorders (mood and anxiety disorders) show a weaker relationship with hospitalization and encephalopathy (Table 1). This may indicate that their occurrence reflects, at least partly, the psychological and other implications of a COVID-19 diagnosis rather than being a direct manifestation of the illness.

HRs for most neurological outcomes were constant, and hence the risks associated with COVID-19 persist until the six-month time point. Longer-term studies will be needed to ascertain the duration of risk, and the trajectory for individual diagnoses.

Our findings are relatively robust, given the sample size, the propensity score matching, and the results of the sensitivity and secondary analyses. Nevertheless, they have weaknesses inherent to an electronic health records study,^28^ such as the unknown completeness of records, lack of validation of diagnoses, and sparse information on socioeconomic and lifestyle factors. These issues primarily affect the incidence estimates rather than hazard ratios, but the choice of cohorts against which to compare COVID-19 outcomes influences the HRs (see Appendix Table S6). The analyses regarding encephalopathy deserve a note of caution. Even amongst hospitalized patients, only about 11% received this diagnosis, even though much higher rates would be expected.^18,29^ Under-recording of delirium and other altered mental states during acute illness is well known, and likely means that the diagnosed cases had prominent and/or sustained features; as such, results for this group should not be generalised to all patients with delirium. Finally, a study of this kind can only demonstrate associations; efforts to identify mechanisms and assess causality will require prospective cohort studies or more elaborate study designs.

## Data Availability

The TriNetX system returned the results of these analyses as csv files which were downloaded and archived. Data presented in this paper and the Appendix can be freely accessed at [URL to be added on publication]. Additionally, TriNetX will grant access to researchers if they have a specific concern (via the third-party agreement option).

## Acknowledgments

PJH and MT were granted unrestricted access to the TriNetX Analytics network for the purposes of research, and with no constraints on the analyses performed nor the decision to publish. Work supported by the National Institute for Health Research (NIHR) Oxford Health Biomedical Research Centre (grant BRC-1215-20005). The views expressed are those of the authors and not necessarily those of the UK National Health Service, NIHR, or the UK Department of Health. MT is an NIHR Academic Clinical Fellow.

