## Appendix for "Six-month Neurological and Psychiatric Outcomes in 236,379 Survivors of COVID-19"

### Supplementary methods

#### TriNetX network

In this section, we provide more details about the TriNetX network, the data source, its advantages and disadvantages. This is largely reproduced from our previous description of the data1.

The data is stored onboard a TriNetX appliance – a physical server residing at the institution's data centre or a virtual hosted appliance. The TriNetX platform is a fleet of these appliances connected into a federated network able to broadcast queries to each appliance. Results are subsequently collected and aggregated.

Once the data is sent to the network, it is mapped to a standard and controlled set of clinical terminologies and undergoes a data quality assessment including 'data cleaning' that rejects records which do not meet the TriNetX quality standards (see next section). HIPAA compliance of the clinical patient data is achieved using de-identification. Different data modalities are available in the network. They include demographics (coded to HL7 version 3 administrative standards), diagnoses (represented by ICD-10-CM codes), procedures (coded in ICD-10-PCS or CPT), and measurements (coded to LOINC). While extensive information is provided about patients' diagnoses and procedures, other variables (such as socioeconomic and lifetime factors are not comprehensively represented).

The data from a typical healthcare organization generally go back around 7 years, with some going back 13 years. The data are continuously updated. Healthcare organisations update their data at various times, with most healthcare organisations refreshing every 1, 2, or 4 weeks. To comply with legal frameworks and ethical guidelines guarding against data re-identification, the identity of participating health-care organisations and their individual contribution to each dataset are not disclosed.

The advantage of EHR data over insurance claim data is that both insured and uninsured patients are included. An advantage of EHR data over survey data is that they represent the diagnostic rates in the population presenting to healthcare facilities. This provides an accurate account of the burden of specific diagnoses on healthcare systems. The downside of relying on diagnoses is that they obviously do not account for undiagnosed patients who might be suffering from the illness but did not seek medical attention. A general limitation of EHR data is that a patient may be seen in different HCOs for different parts of their care and if one HCO is not part of the federated network then part of their medical records may not be available. Using a network of HCOs (rather than a single HCO) limits this possibility but does not fully remove it.

#### Definition of cohorts

The two control cohorts used consisted of patients with a diagnosis of influenza (ICD-10 codes J09-J11) and patients with another respiratory tract infection (ICD-10 codes J00-J06 'Acute upper respiratory tract infections', J09-J18 'Influenza and pneumonia', and J20-J22 'other acute lower respiratory tract infection').

Because some patients with the control index event might have had COVID-19 at a different point in time, we excluded from the control cohorts all those who had COVID-19 at any point in time. To avoid any contamination between cohorts, COVID-19 as an exclusion criterion was defined in the broader sense to be all patients with a confirmed diagnosis of COVID-19 (ICD-10 code U07.1) but also patients with an unconfirmed COVID-19 diagnosis (U07.2), a recorded positive PCR test for COVID-19, or any of the following recorded on or after January 20, 2020: Pneumonia due to SARS-associated coronavirus (J12.81), Other coronavirus as the cause of disease classified elsewhere (B97.29), or Coronavirus infection unspecified (B34.2). Inclusion of the latter three diagnostic codes captures patients who receive a COVID-19 diagnosis in the early stage of the pandemic when the ICD code for COVID-19 (U07) was not yet defined.

**Definition of covariates**

To reduce the effect of confounding on associations between a diagnosis of COVID-19 and a subsequent neurologic or psychiatric diagnosis, cohorts were matched for established or suspected risk factors for COVID-19[2–5] and for established risk factors for COVID-19 death[6] (taken to be risk factors of a more severe COVID-19 illness). The following confounding factors were therefore included (with ICD-19/CDC codes in brackets):

1) **Age** at the time of diagnosis
2) **Sex** coded as female, male, or other.
3) **Race** encoded as 6 separate dichotomous variables: White (2106-3), Black or African American (2054-5), American Indian or Alaska Native (1002-5), Asian (2028-9), Native Hawaiian or Other Pacific Islander (2076-8), or Unknown Race (2131-1)
4) **Ethnicity** encoded as Hispanic or Latino (2135-2), Not Hispanic or Latino (2186-5), or Unknown Ethnicity
5) **Socioeconomic deprivation** encoded as the ICD-10 code for Problems related to housing and economic circumstances (Z59)
6) **Obesity** encoded as one dichotomous variable and one categorical variable: Overweight and obesity (E66) and body mass index (categorised into < 25 kg/m$^2$, 25-30 kg/m$^2$, ≥ 30 kg/m$^2$)
7) **Hypertension** encoded as 2 dichotomous and 2 categorical variables: Hypertensive diseases (I10-I16), the now deprecated version that was used until 2018 Hypertension diseases (I10-I15), measurements of systolic blood pressure (categorised into < 140mmHg, 140-160mmHg, and ≥ 160mmHg), and diastolic blood pressure (categorised into < 90mmHg, 90-100mmHg, and ≥ 100mmHg)
8) **Diabetes mellitus** encoded as 2 dichotomous variables: Type 1 diabetes mellitus (E10) and Type 2 diabetes mellitus (E11)
9) **Chronic lower respiratory diseases** encoded by each sub-category of the corresponding ICD-10 group: Bronchitis, not specified as acute or chronic (J40), Simple and mucopurulent chronic bronchitis (J41), Unspecified chronic bronchitis (J42), Emphysema (J43), Other chronic obstructive pulmonary disease (J44), Asthma (J45), Bronchiectasis (J47)
10) **Nicotine dependence** encoded as the corresponding ICD-10 diagnosis (F17.2)
11) **Substance use disorder** encoded as the ICD-10 code for mental and behavioral disorders due to psychoactive substance use (F10-F19)
12) **Heart diseases** encoded as 2 categorical variables: Ischaemic heart disease (I20-I25) and Other forms of heart disease (I30-I52)
13) **Chronic kidney disease** encoded as 2 dichotomous variables: Chronic kidney disease (N18) and Hypertensive chronic kidney disease (I12)
14) **Chronic liver disease** encoded as 8 categorical variables: Alcoholic liver disease (K70), Hepatic failure, not elsewhere classified (K72), Chronic hepatitis, not elsewhere classified (K73), Fibrosis and cirrhosis of liver (K74), Fatty (change of) liver, not elsewhere classified (K76.0), Chronic passive congestion of liver (K76.1), Portal hypertension (K76.6), Other specified diseases of liver (K76.8)
15) **Stroke** encoded as the dichotomous variable Cerebral infarction (I63)
16) **Dementia** encoded as 6 dichotomous variables: Vascular dementia (F01), Dementia in other diseases classified elsewhere (F02), Unspecified dementia (F03), Alzheimer's disease (G30), Frontotemporal dementia (G31.0), and Dementia with Lewy bodies (G31.83)
17) **Cancer and haematological cancer in particular** encoded as 2 dichotomous variables: Neoplasms (C00-D49) and Malignant neoplasms of lymphoid, hematopoietic and related tissue (C81-C96)
18) **Organ transplant** encoded as 2 dichotomous variables: Renal Transplantation Procedures and Liver Transplantation Procedures
19) **Rheumatoid arthritis** encoded as 2 dichotomous variables: Rheumatoid arthritis with rheumatoid factor (M05) and Other rheumatoid arthritis (M06)
20) **Lupus** encoded as a dichotomous variable corresponding ICD-10 code (M32)
21) **Psoriasis** encoded as a dichotomous variable corresponding ICD-10 code (L40)
22) **Other immunosuppression** encoded as a dichotomous variable “Certain disorders involving the immune mechanism” (D80-D89)

Each individual code was considered a confounding factor in and of itself so that matching was achieved for each of them individually. For instance, matching was achieved for each subcategory (and not just for the whole category) of chronic lower respiratory diseases. For variables representing diagnoses and socioeconomic deprivation, an individual was considered positive if the diagnostic was recorded at least once in their health record before the index event. For categorical variables representing measurements (i.e. BMI and blood pressures), all available measurements for all individuals were used and propensity score matching sought to define cohorts with similar numbers of measurements falling into each category.

**Definition of outcomes**

All outcomes were defined as a diagnosis recorded in the patient's electronic health record between 1 day and 180 days after the index event. As mentioned in the manuscript, for chronic illnesses, only *first* diagnoses were counted (i.e. patients with the diagnosis before the index event were excluded from the survival analysis). For diagnoses that can recur or relapse, we separately estimated the incidence of *first* diagnosis and the incidence of *any* diagnoses. For all other diagnoses, we estimated the incidence of *any* diagnoses.

Specifically the following ICD-10 codes (with the ICD-10 labels in brackets) were used to define outcomes:

1) Intracranial haemorrhage (first and any diagnosis): I60 (non-traumatic subarachnoid haemorrhage), I61 (non-traumatic intracerebral haemorrhage), and I62 (other and unspecified non-traumatic intracranial haemorrhage)
2) Ischaemic stroke (first and any diagnosis): I63 (cerebral infarction)
3) Parkinsonism (first diagnosis): G20 (Parkinson's disease) or G21 (Secondary parkinsonism)
4) Guillain-Barre syndrome (any diagnosis): G61.0 (Guillain-Barre syndrome)
5) Nerve/nerve root/plexus disorders (any diagnosis): G50-G59 (Nerve, nerve root and plexus disorders)
6) Myoneural junction/muscle disease (first diagnosis): G70-G73 (Diseases of myoneural junction and muscle)
7) Encephalitis (any diagnosis): G04 (Encephalitis, myelitis and encephalomyelitis), G05 (Encephalitis, myelitis and encephalomyelitis in diseases classified elsewhere), A86 (Unspecified viral encephalitis), or A85.8 (Other specified viral encephalitis)
8) Dementia (first diagnosis): F01 (Vascular dementia), F02 (Dementia in other diseases classified elsewhere), F03 (Unspecified dementia), G30 (Alzheimer's disease), G31.0 (Frontotemporal dementia), G31.83 (Dementia with Lewy bodies)
9) Mood/Anxiety/Psychotic disorder (first and any diagnosis): F20-F29 (Schizophrenia, schizotypal, delusional, and other non-mood psychotic disorders), F30-F39 (Mood disorders), F40-F48 (Anxiety, dissociative, stress-related, somatoform and other nonpsychotic mental disorders)
    a) Mood disorder (first and any diagnosis): F30-F39
    b) Anxiety disorder (first and any diagnosis): F40-F48
    c) Psychotic disorder (first and any diagnosis): F20-F29
10) Substance misuse (first and any diagnosis): F10-F19 (Mental and behavioral disorders due to psychoactive substance use)
11) Insomnia (first and any diagnosis): F51.0 (Insomnia not due to a substance or known physiological condition) or G47.0 (Insomnia)

In the definition of mood disorder and substance misuse, ICD-10 codes representing remission (e.g. F32.4 - Major depressive disorder, single episode, in partial remission) were excluded.

**Details on secondary analyses**

Encephalopathy was defined as the presence of any of the following diagnostic code between 4 days before and 2 weeks after the COVID-19 diagnosis: Other and unspecified encephalopathy (ICD-10 code G93.4), Delirium (F05), Other mental disorders due to known physiological condition (F06), Personality and behavioral disorders due to known physiological condition (F07), Disorientation (R41.0), Somnolence (R40.0), and Stupor (R40.1). The combination of these diagnostic codes aimed to capture various clinical presentations of encephalopathy which can all represent a change from baseline cognitive status[7]. Furthermore, they account for semantic differences across disciplines. Our choice of the term 'encephalopathy' to characterise this cohort reflects both the fact that other codes are clinical manifestations of encephalopathy the fact that encephalopathy (G93.4) was the most prevalent code used in this cohort (present in 66.0% of patients).

To provide benchmarks for the incidence of neurologic and psychiatric sequelae of health events, we compared these incidences between the COVID-19 cohort and 4 other matched cohorts of patients defined by another index event:

- Skin infection: ICD-10 codes L00-L08 ('Infection of the skin and subcutaneous tissue')
- Urolithiasis: ICD-10 codes N20-N23
- Fracture of a large bone: ICD-10 codes S32 ('Fracture of lumbar spine and pelvis'), S42 ('Fracture of shoulder and upper arm'), S52 ('Fracture of forearm'), S72 ('Fracture of femur'), S82 ('Fracture of lower leg including ankle')
- Pulmonary embolism: ICD-10 code I26.

This analysis was achieved using the same survival analysis as in the primary analysis and by only altering the index event of the control cohort. These additional control index events were selected as they represent a broad range of common acute presentations with a range of anticipated neurologic and psychiatric sequelae.

**Details on statistical analyses**

In propensity score matching, the propensity score was calculated using a logistic regression (implemented by the function LogisticRegression of the scikit-learn package in Python 3.7) including each of the covariates mentioned above. To eliminate the influence of ordering of records, the order of the records in the covariate matrix were randomised before matching.

The assumption that the hazards were proportional when accounting for the two phases was tested using the generalized Schoenfeld approach implemented in the cox.zph function of the survival package (version 3.2.3) in R. If the proportional hazard assumption was found to be violated, then the time-varying HR was assessed using natural cubic splines (in log-time) to the log-cumulative hazard[8]. This was achieved using the generalized survival models of the rstpm2 package (version 1.5.1) in R[9]. As recommended by Royston and Parmar[8], splines with 1, 2, and 3 degrees of freedom were estimated for both the baseline log-cumulative hazard and its cohort dependency and the number of degrees of freedom leading to the lowest Akaike Information Criterion (AIC) was selected. This was achieved on a per-comparison basis so that more complex time dependency (i.e. higher number of degrees of freedom) could be selected for a specific comparison if there was enough evidence in the data to support such complexity.

#### Supplementary figures

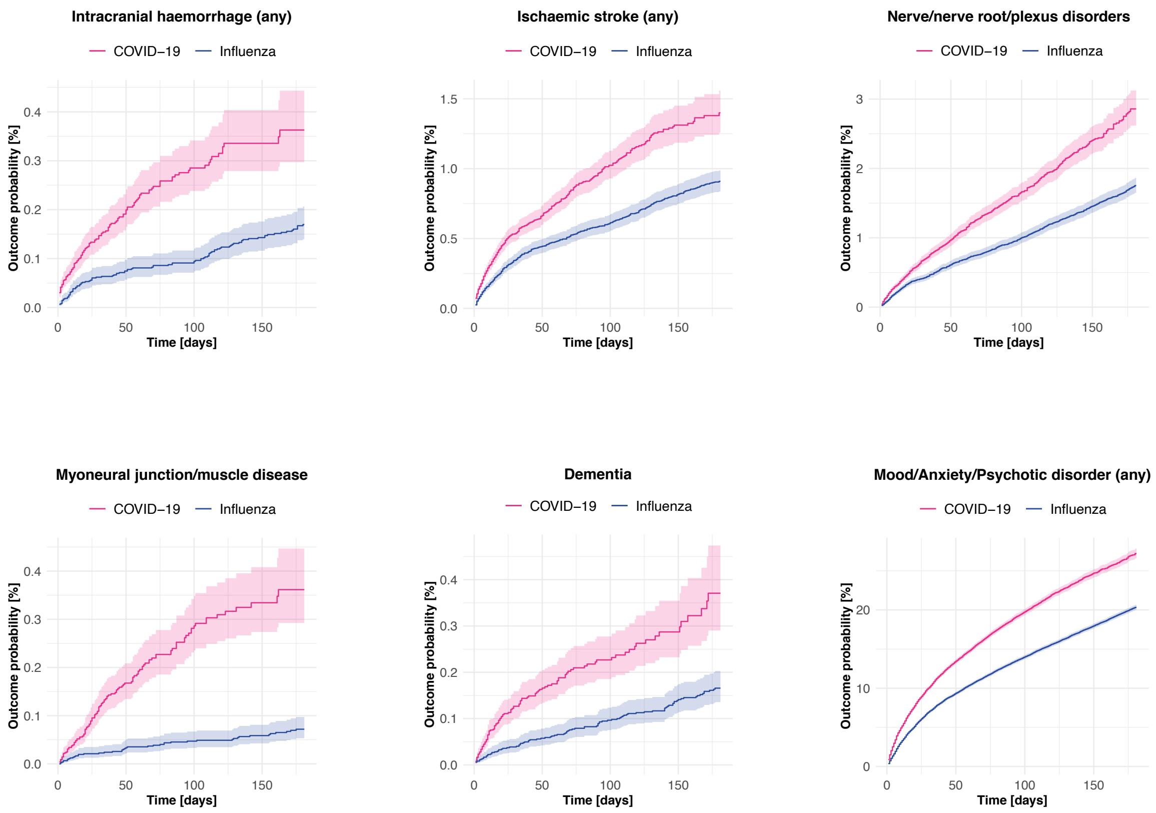


**Figure S1 –** Kaplan-Meier curves for the comparison between the cohort of patients with COVID-19 and a matched cohort of patients with influenza for the same outcomes as in Figure 1 of the main manuscript. Shaded areas represent 95% confidence intervals.

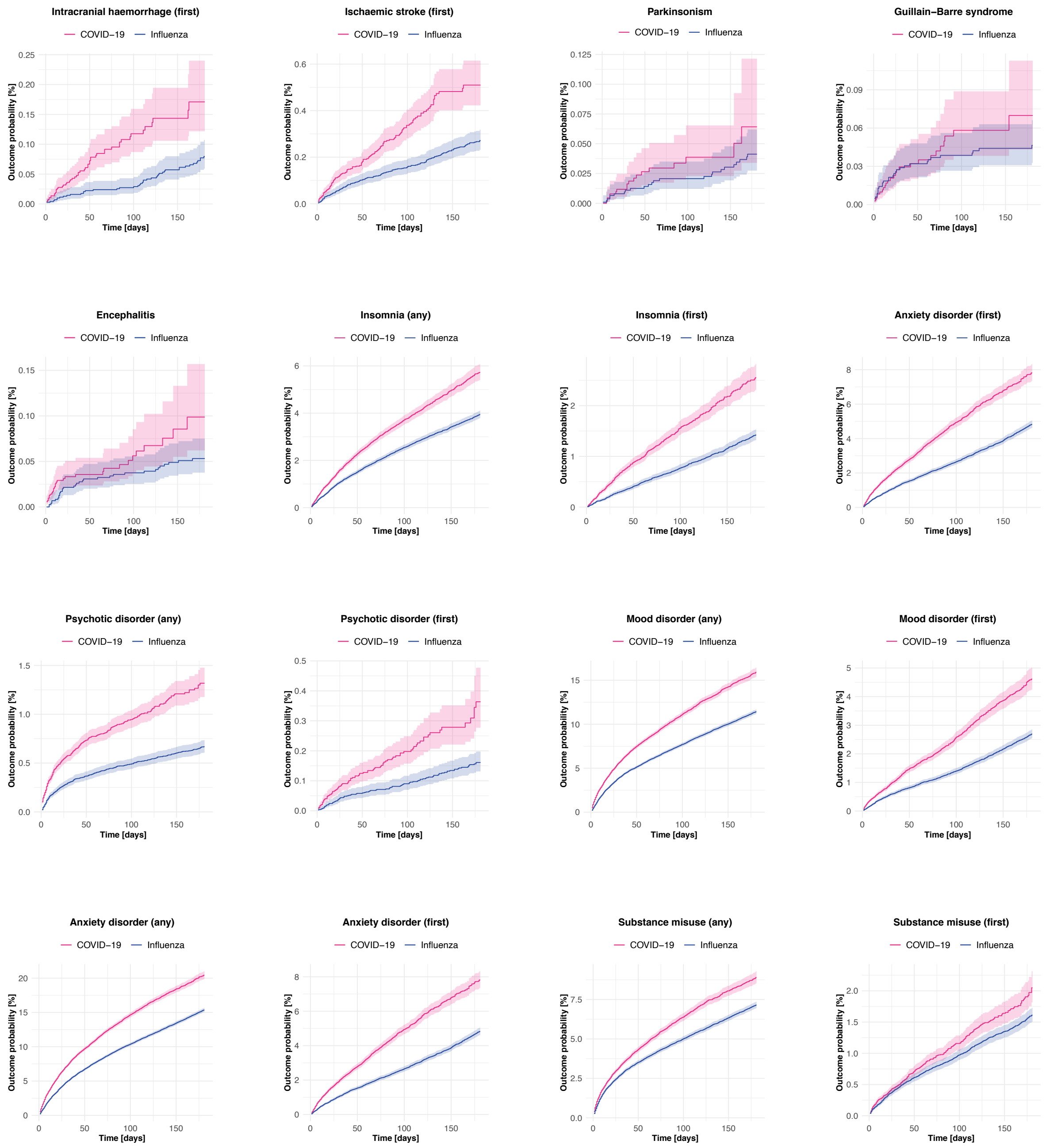


**Figure S2 –** Kaplan-Meier curves for the comparison between the cohort of patients with COVID-19 and a matched cohort of patients with influenza for the outcomes not included in Figure S1. Shaded areas represent 95% confidence intervals.

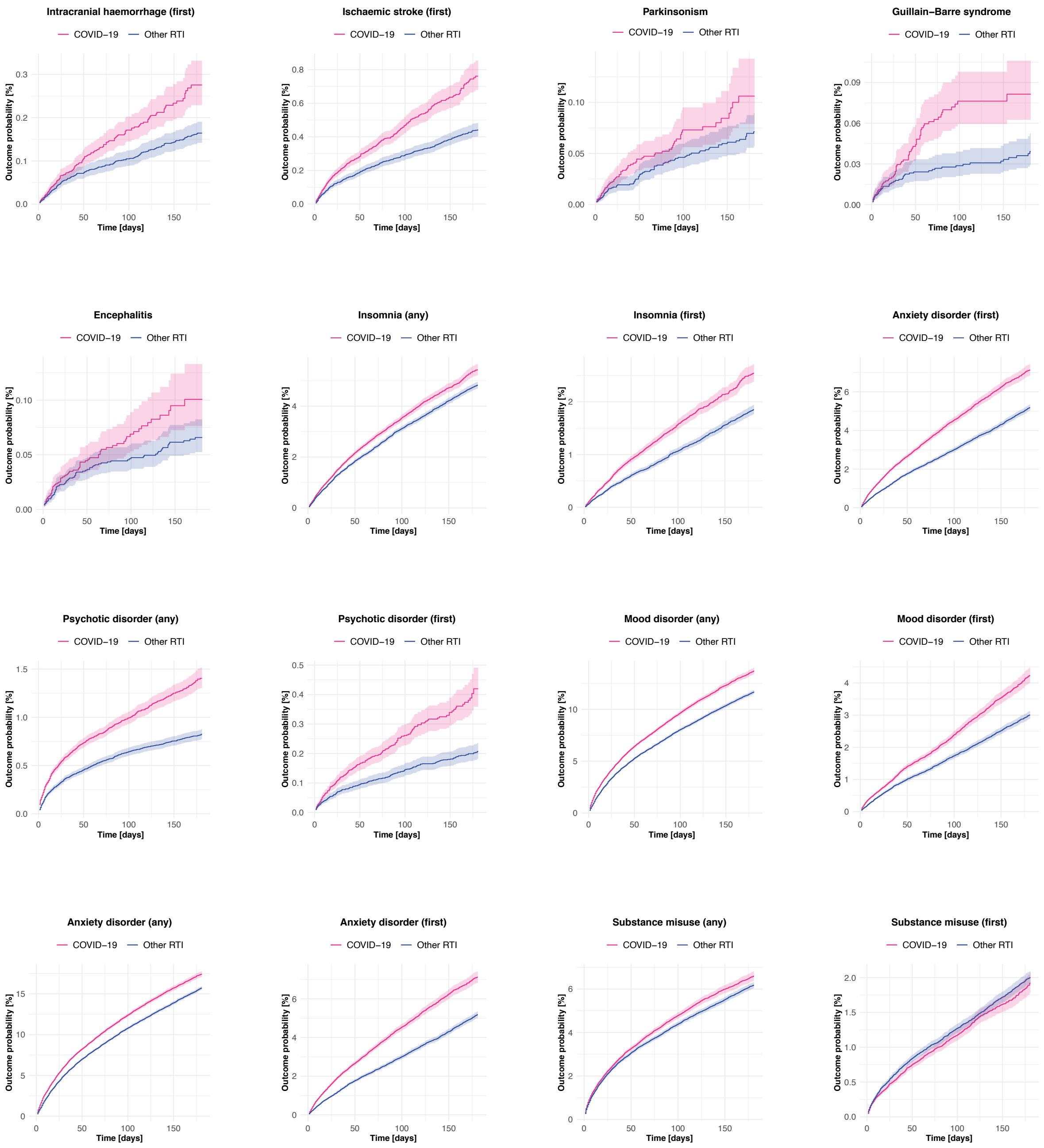


**Figure S3 –** Kaplan-Meier curves for the comparison between the cohort of patients with COVID-19 and a matched cohort of patients with other respiratory tract infections for the outcomes not included in Figure S1. Shaded areas represent 95% confidence intervals.

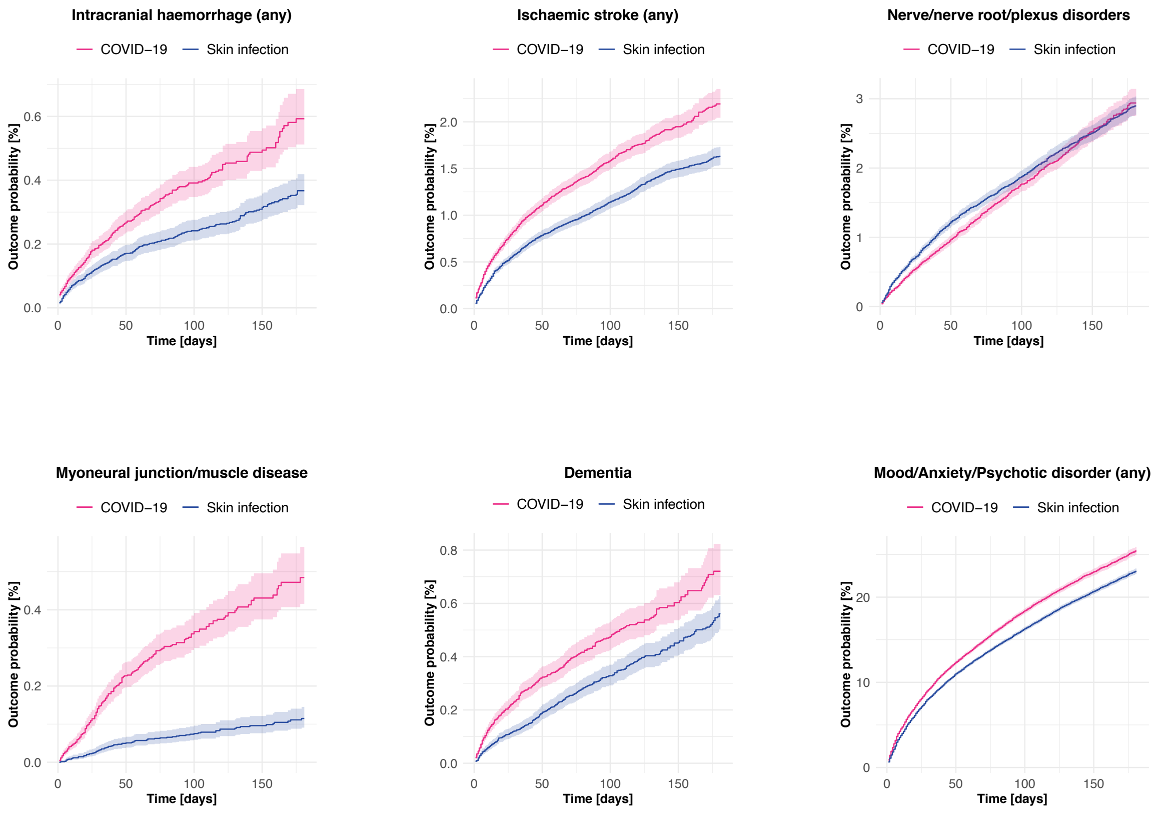


**Figure S4 –** Kaplan-Meier curves for the comparison between the cohort of patients with COVID-19 and a matched cohort of patients with skin infection for the same outcomes as in Figure 1 of the main manuscript. Shaded areas represent 95% confidence intervals.

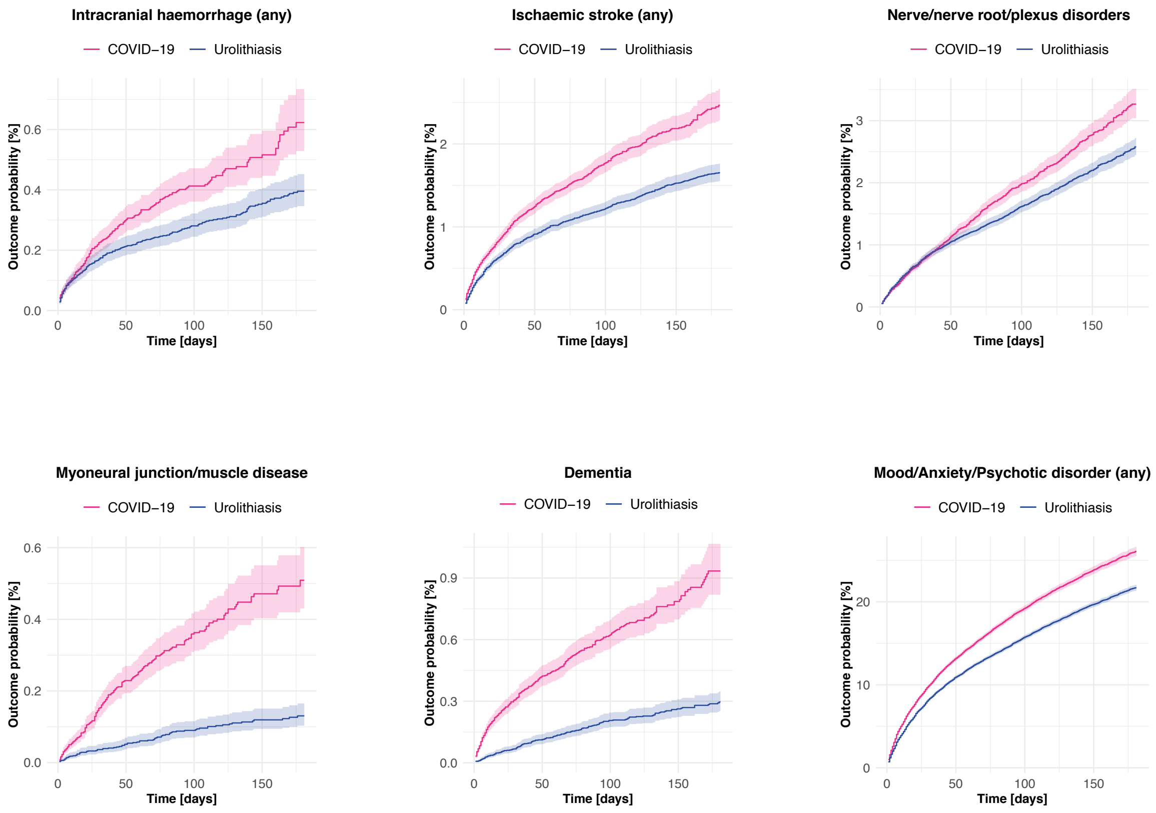


**Figure S5 –** Kaplan-Meier curves for the comparison between the cohort of patients with COVID-19 and a matched cohort of patients with urolithiasis for the same outcomes as in Figure 1 of the main manuscript. Shaded areas represent 95% confidence intervals.

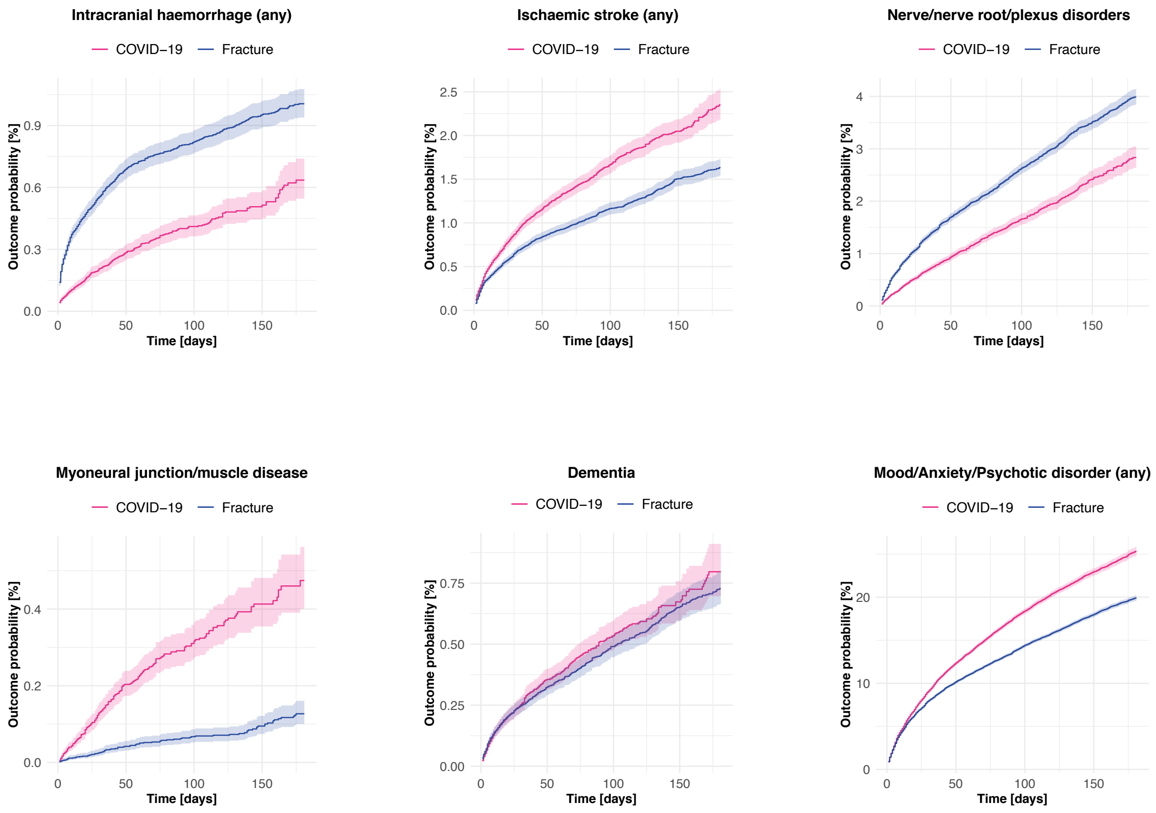


**Figure S6 –** Kaplan-Meier curves for the comparison between the cohort of patients with COVID-19 and a matched cohort of patients with fracture of a large bone for the same outcomes as in Figure 1 of the main manuscript. Shaded areas represent 95% confidence intervals.

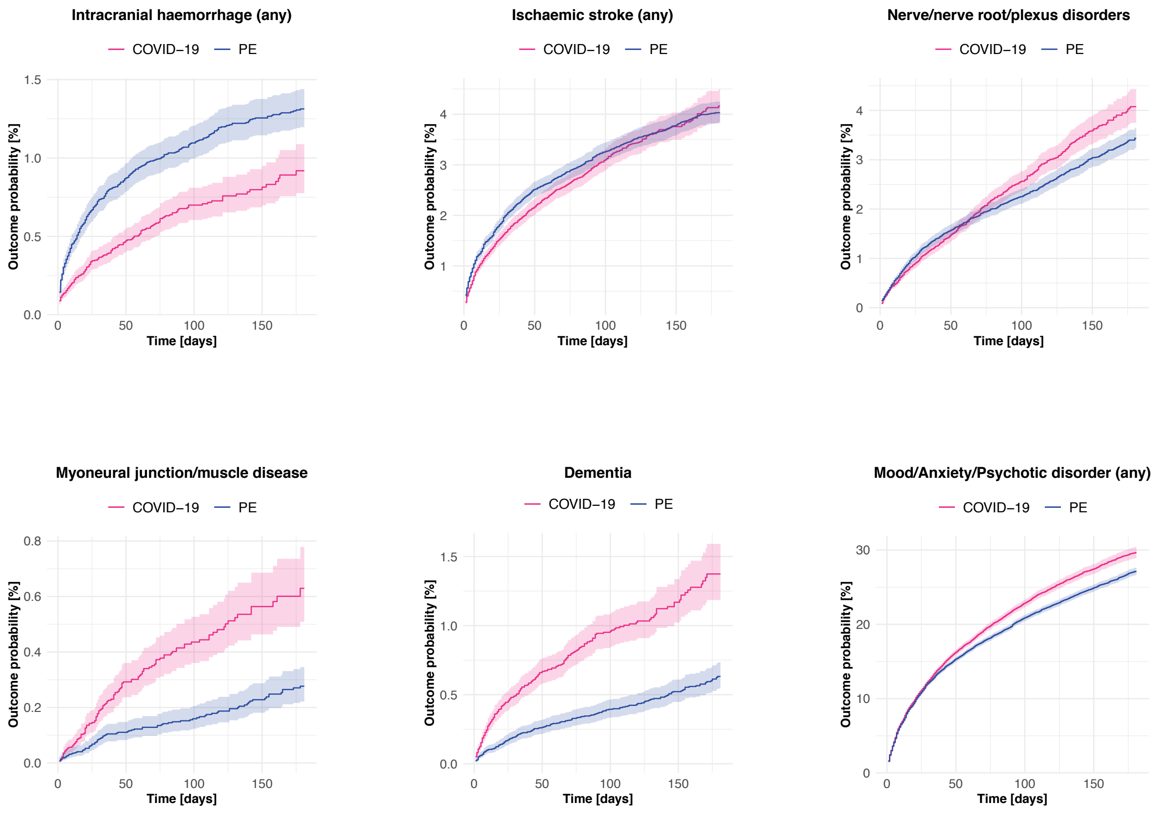


**Figure S7 –** Kaplan-Meier curves for the comparison between the cohort of patients with COVID-19 and a matched cohort of patients with pulmonary embolism (PE) for the same outcomes as in Figure 1 of the main manuscript. Shaded areas represent 95% confidence intervals.

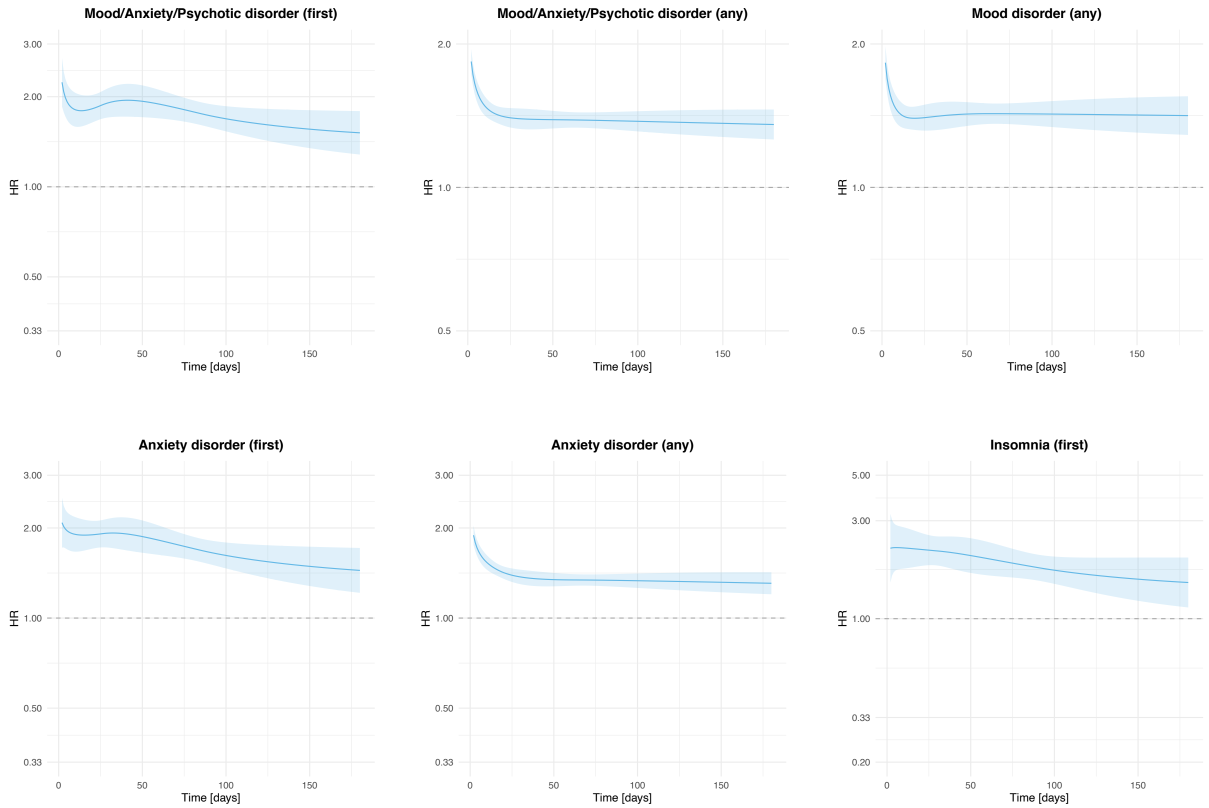


**Figure S8** – Time-varying hazard ratios for the outcomes for which there was evidence of non-proportionality of hazards in the main comparison between the cohort of patients with COVID-19 and a matched cohort of patients with influenza. Shaded area represents a 95% confidence interval.

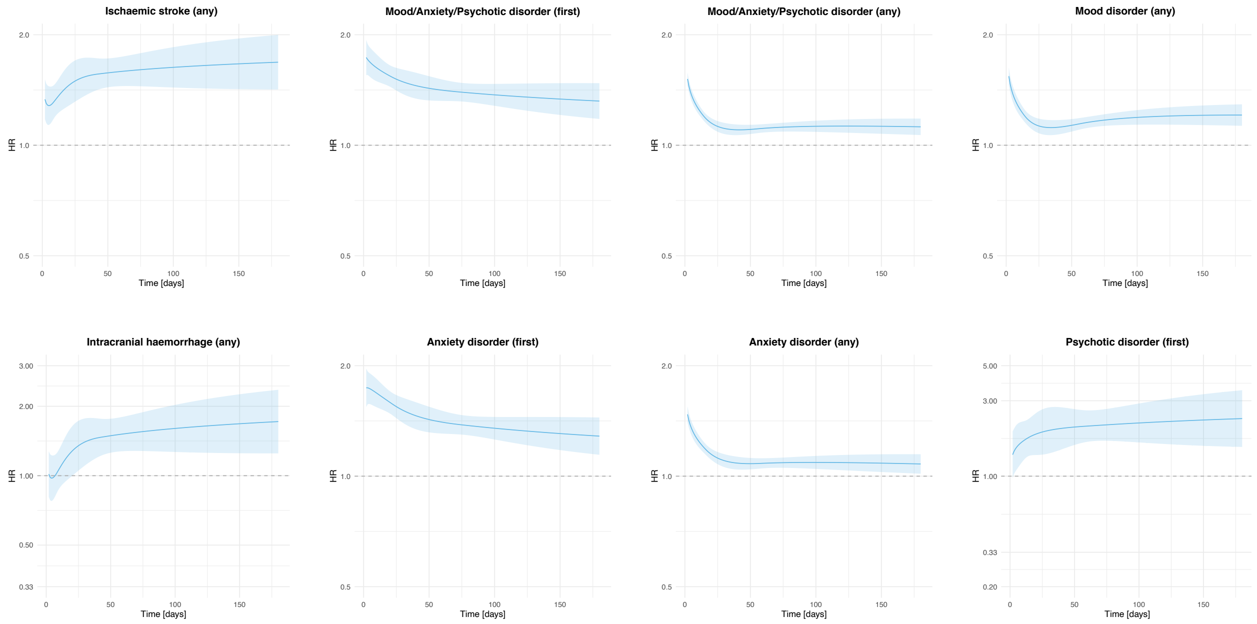


**Figure S9** – Time-varying hazard ratios for the outcomes for which there was evidence of non-proportionality of hazards in the main comparison between the cohort of patients with COVID-19 and a matched cohort of patients with other respiratory tract infections. Shaded area represents a 95% confidence interval.

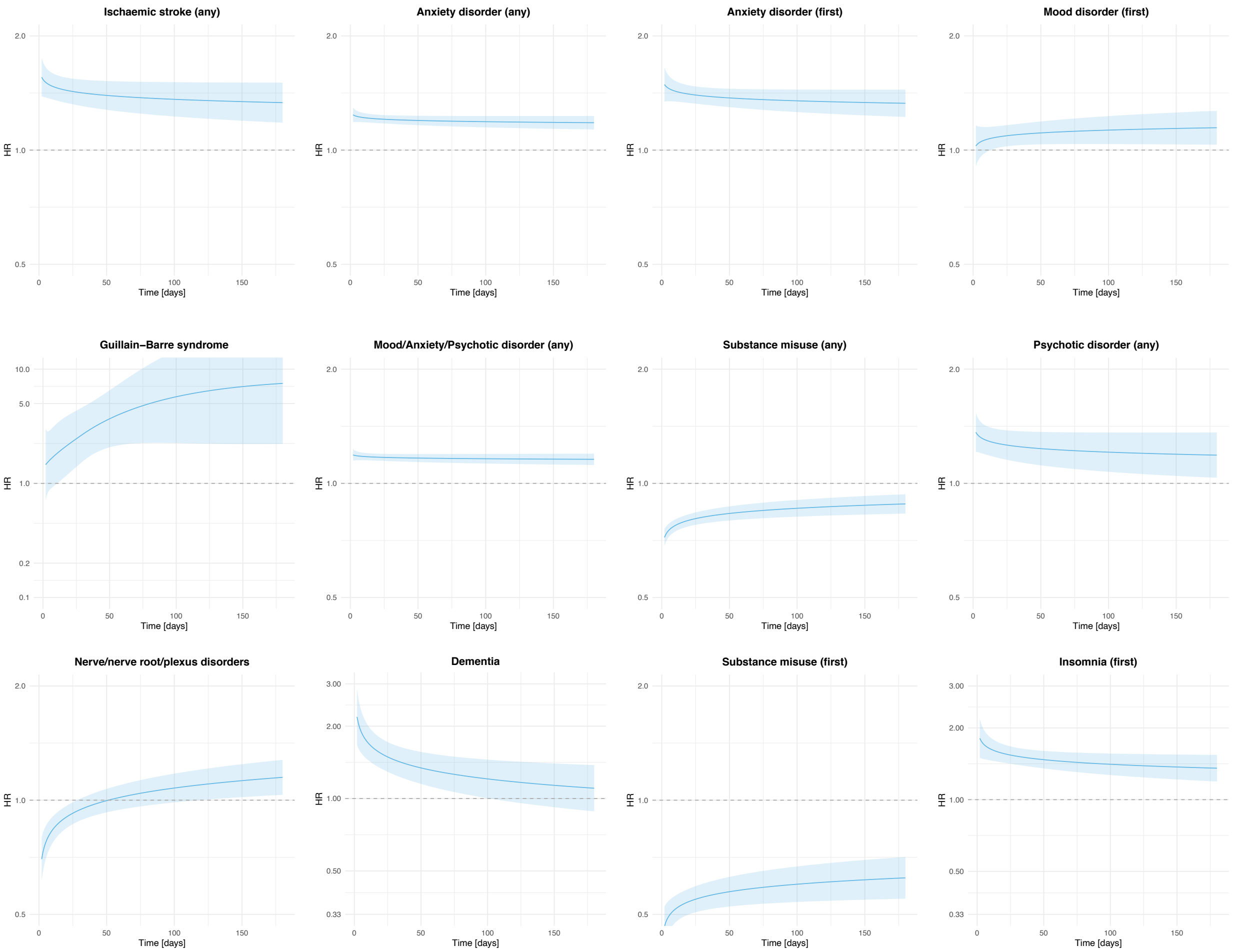


**Figure S10** – Time-varying hazard ratios for the outcomes for which there was evidence of non-proportionality of hazards in the comparison between the cohort of patients with COVID-19 and a matched cohort of patients with skin infection. Shaded area represents a 95% confidence interval.

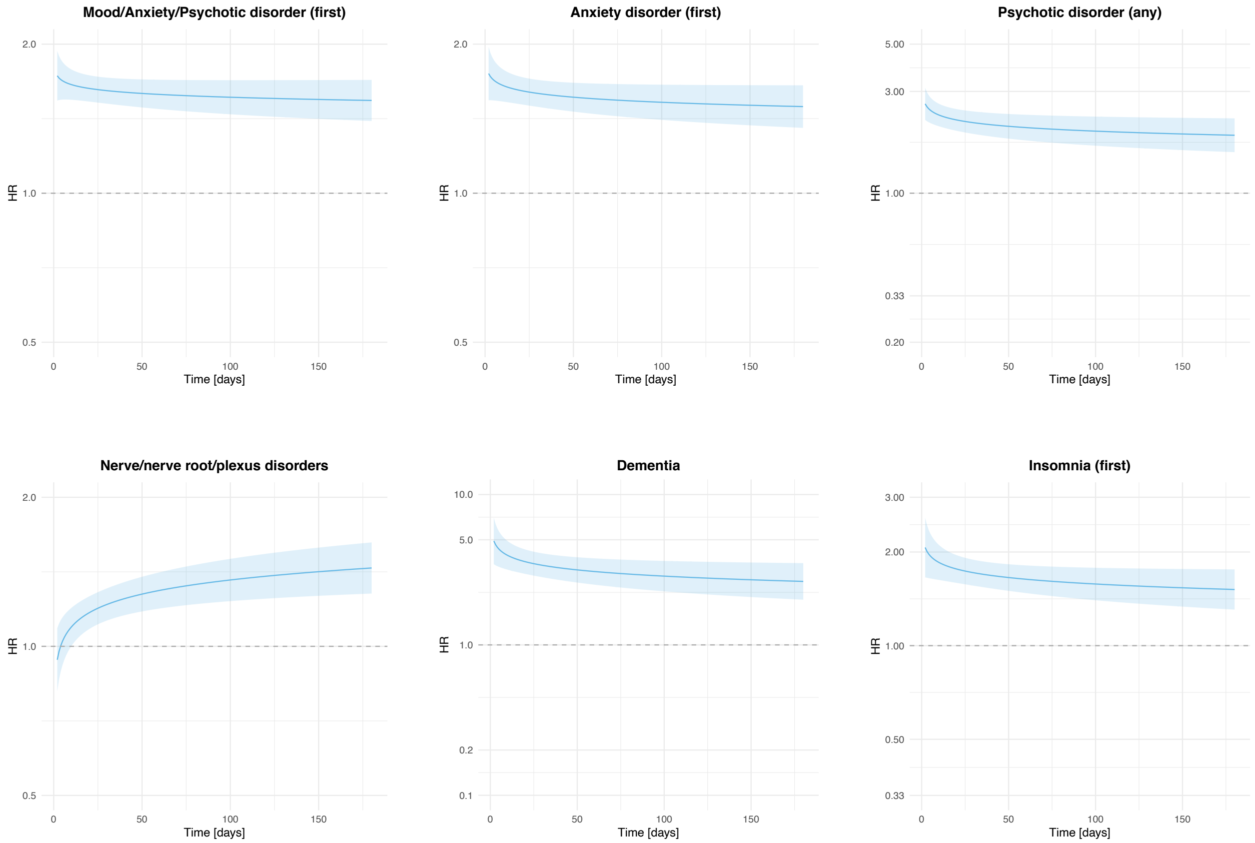


**Figure S11** – Time-varying hazard ratios for the outcomes for which there was evidence of non-proportionality of hazards in the comparison between the cohort of patients with COVID-19 and a matched cohort of patients with urolithiasis. Shaded area represents a 95% confidence interval.

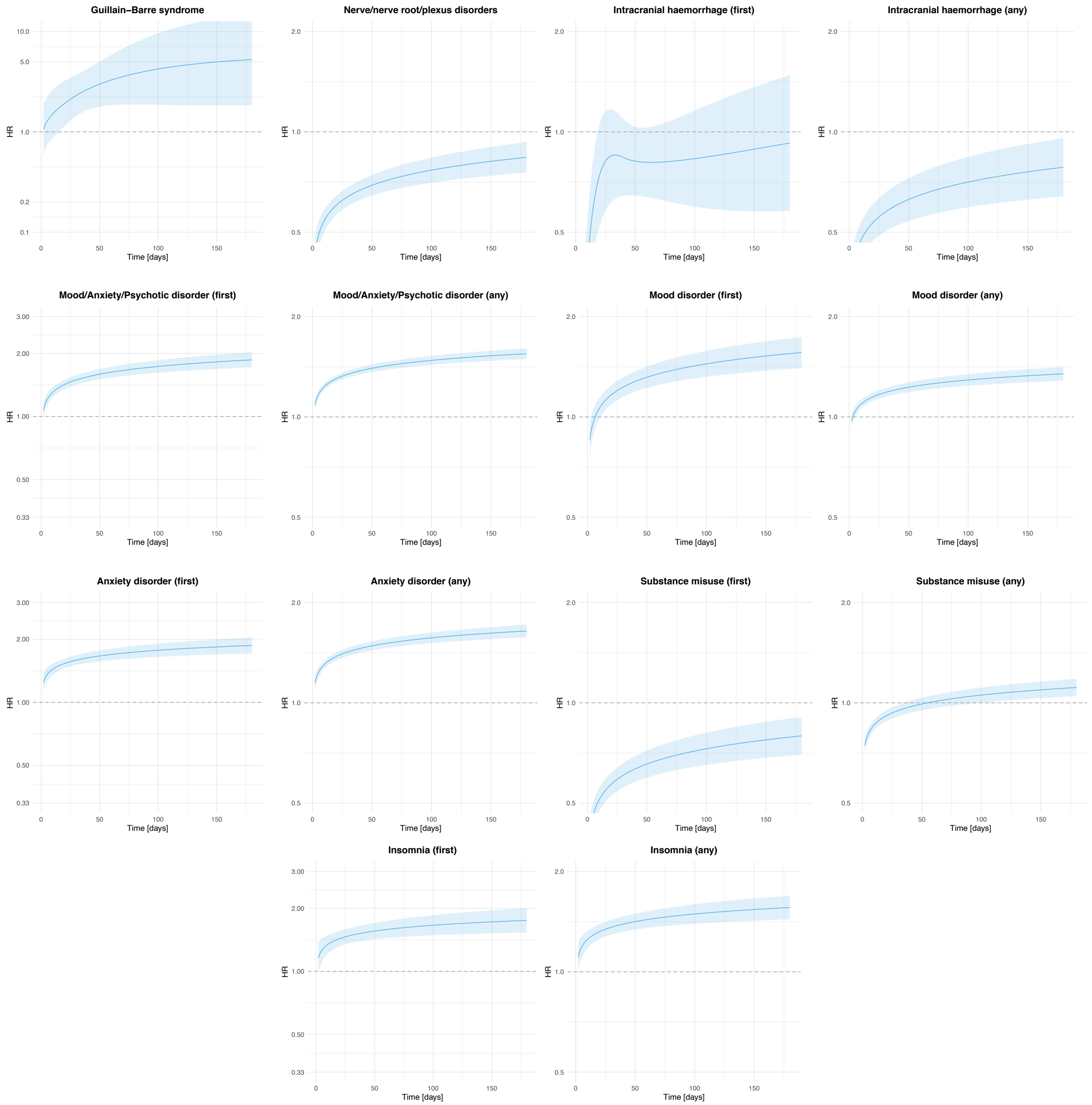


**Figure S12** – Time-varying hazard ratios for the outcomes for which there was evidence of non-proportionality of hazards in the comparison between the cohort of patients with COVID-19 and a matched cohort of patients with fracture of a large bone. Shaded area represents a 95% confidence interval.

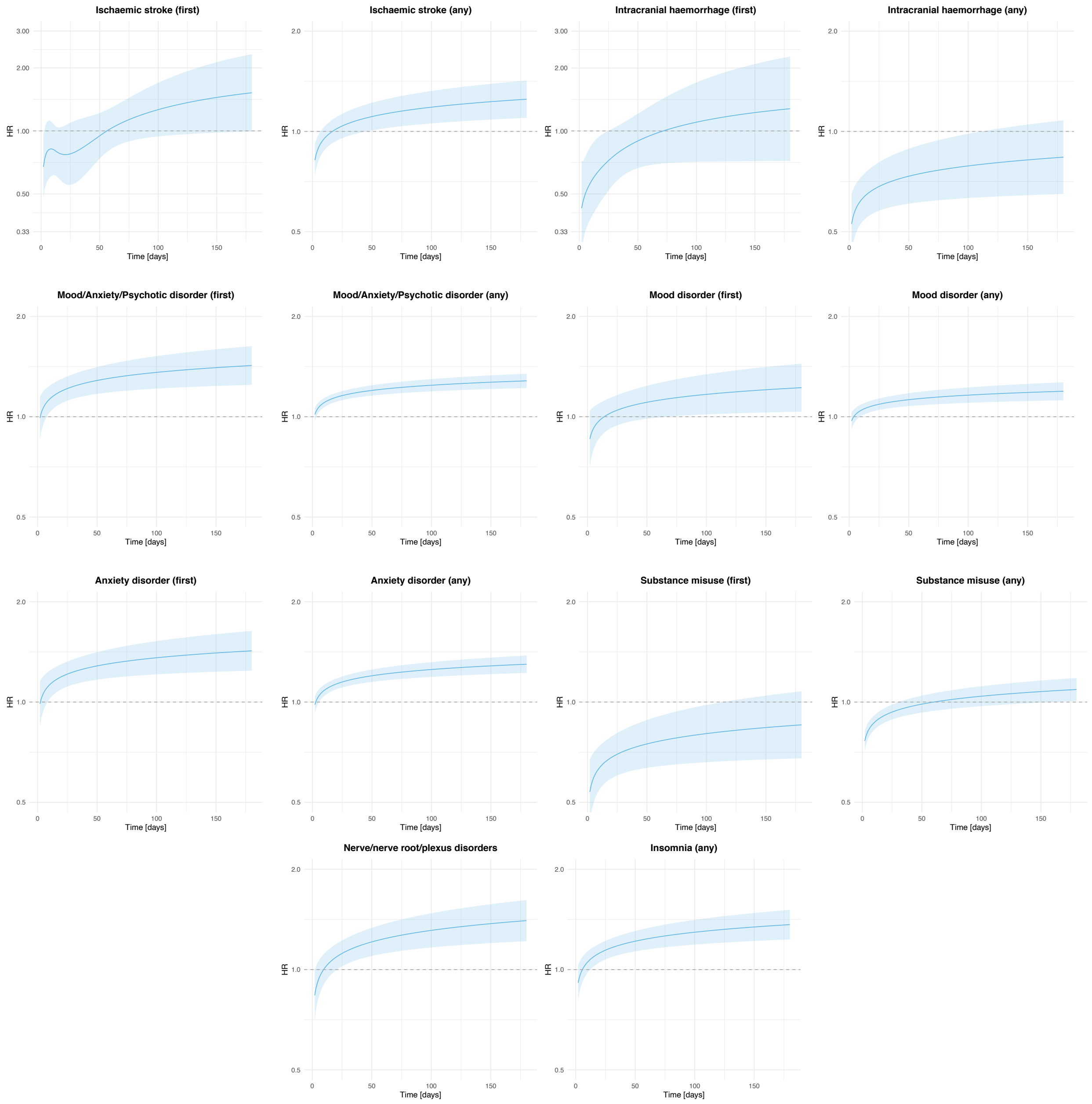


**Figure S13** – Time-varying hazard ratios for the outcomes for which there was evidence of non-proportionality of hazards in the comparison between the cohort of patients with COVID-19 and a matched cohort of patients with pulmonary embolism. Shaded area represents a 95% confidence interval.

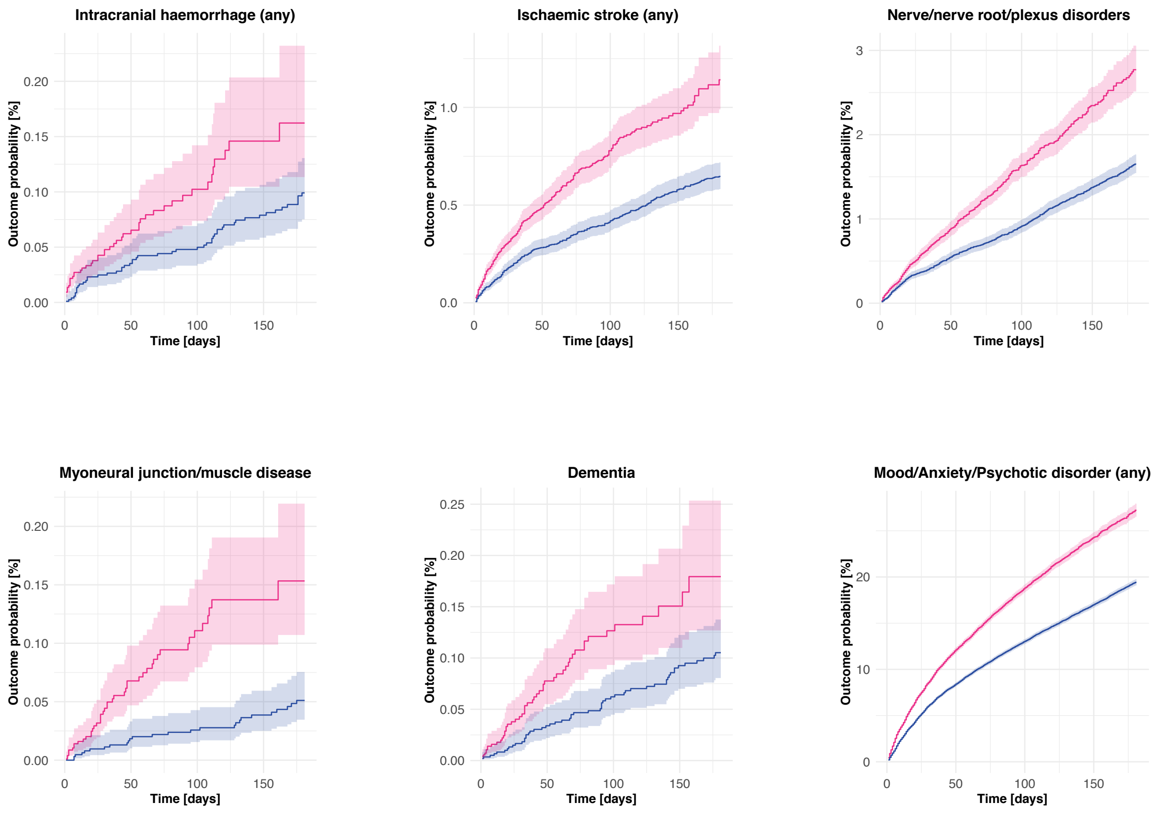


**Figure S14 –** Kaplan-Meier curves for the comparison between the cohort of patients with COVID-19 who did not require hospitalization (red) and a matched cohort of patients with influenza who did not require hospitalization (blue) for the same outcomes as in Figure 1 of the main manuscript. Shaded areas represent 95% confidence intervals.

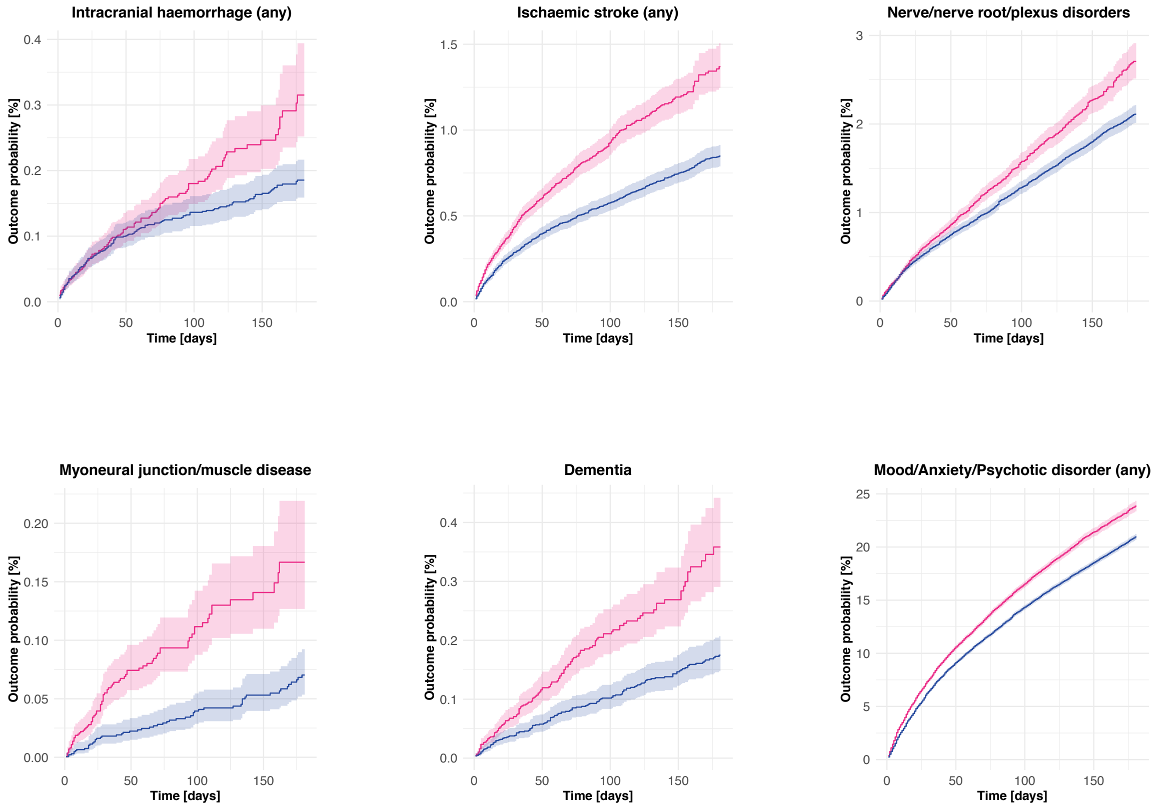


**Figure S15 –** Kaplan-Meier curves for the comparison between the cohort of patients with COVID-19 who did not require hospitalization (red) and a matched cohort of patients with other respiratory tract infections who did not require hospitalization (blue) for the same outcomes as in Figure 1 of the main manuscript. Shaded areas represent 95% confidence intervals.

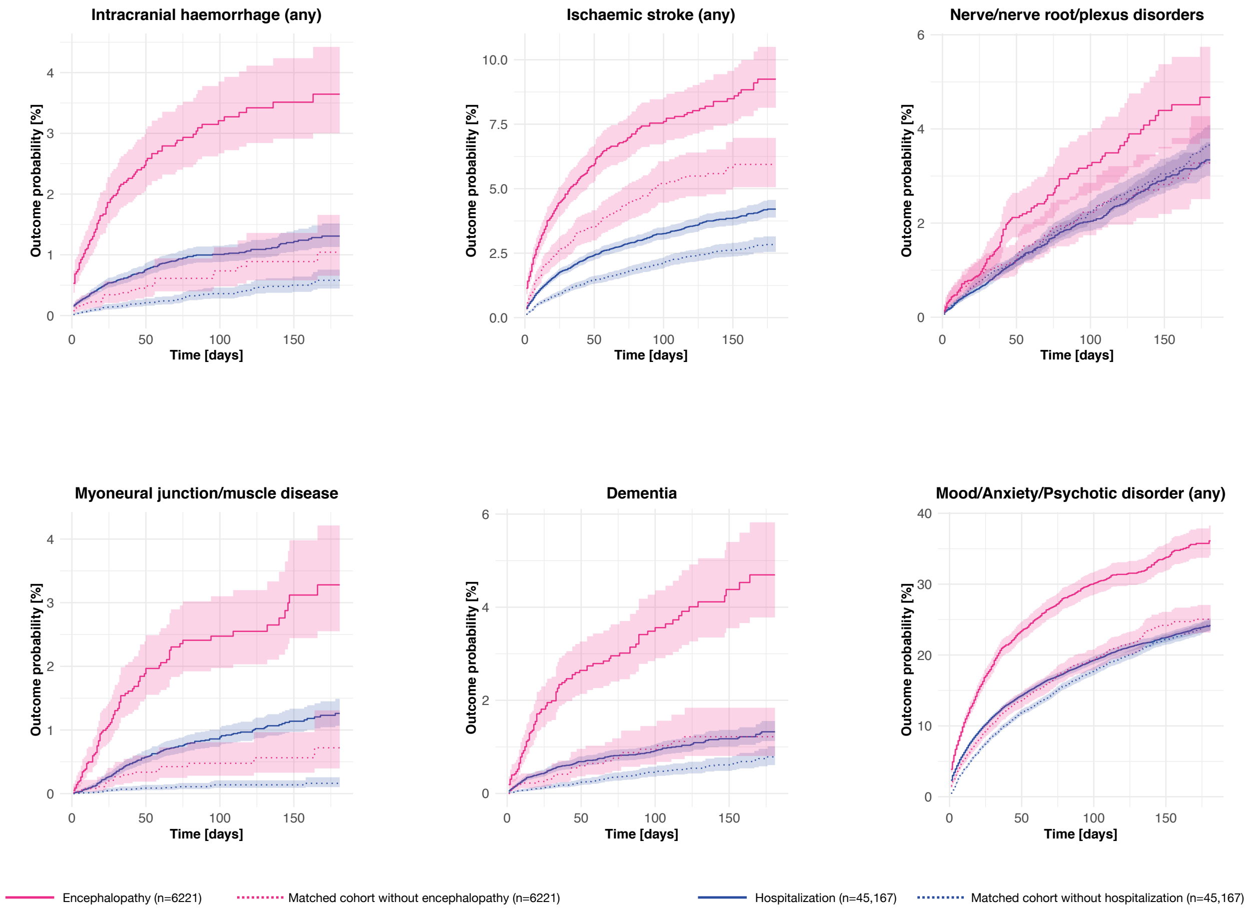


**Figure S16 –** Same figure as Figure 2 of the main manuscript but with 95% confidence intervals displayed

#### Supplementary tables

**Table S1** – All baseline characteristics used in the propensity score matching. In addition, the incidence of neurologic or psychiatric sequela except dementia is presented. This is informative because patients who had a diagnosis of dementia (e.g. Unspecified dementia) before their diagnosis of COVID-19 and then go on to have another dementia diagnosis (e.g. Vascular dementia) would count towards the incidence of any diagnosis (alongside those for whom this would be their first diagnosis of dementia). The incidence of 'any first' diagnosis (presented in Table 1 of the main manuscript) is not affected by this limitation since it excludes all previous diagnoses of dementia.

| | **All patients** | **Non-hospitalized patients** | **Hospitalized patients** | **Patients with encephalopathy** |
|---|---|---|---|---|
| Cohort size, n (%) | 236379 | 190077 | 46302 | 6229 |
| DEMOGRAPHICS | | | | |
| **Age**, mean (SD), y | 46 (19.7) | 43.3 (19.0) | 57 (18.7) | 66.7 (17.0) |
| Sex, n (%) | | | | |
| Female | 131460 (55.6) | 107730 (56.7) | 23730 (51.2) | 2909 (46.7) |
| Male | 104015 (44.0) | 81512 (42.9) | 22503 (48.6) | 3307 (53.1) |
| Other | 904 (0.4) | 835 (0.4) | 69 (0.1) | 13 (0.2) |
| Race, n (%) | | | | |
| White | 135143 (57.2) | 109635 (57.7) | 25508 (55.1) | 3331 (53.5) |
| Black or African American | 44459 (18.8) | 33868 (17.8) | 10591 (22.9) | 1552 (24.9) |
| Asian | 6979 (3.0) | 5432 (2.9) | 1547 (3.3) | 243 (3.9) |
| American Indian or Alaska Native | 962 (0.4) | 716 (0.4) | 246 (0.5) | 21 (0.3) |
| Native Hawaiian or Other Pacific Islander | 751 (0.3) | 585 (0.3) | 166 (0.4) | 11 (0.2) |
| Unknown | 48085 (20.3) | 39841 (21.0) | 8244 (17.8) | 1071 (17.2) |
| Ethnicity, n (%) | | | | |
| Hispanic or Latino | 37772 (16.0) | 29155 (15.3) | 8617 (18.6) | 895 (14.4) |
| Not Hispanic of Latino | 134075 (56.7) | 106844 (56.2) | 27231 (58.8) | 3873 (62.2) |
| Unknown | 64532 (27.3) | 54078 (28.5) | 10454 (22.6) | 1461 (23.5) |
| Problems related to housing and economic circumstances, n (%) | 2452 (1.04) | 1400 (0.7) | 1052 (2.27) | 216 (3.5) |
| COMORBIDITIES, n (%) | | | | |
| Overweight and obesity | 42871 (18.1) | 30198 (15.9) | 12673 (27.4) | 1838 (29.5) |
| Hypertensive disease | 71014 (30.0) | 47516 (25.0) | 23498 (50.7) | 4591 (73.7) |
| Diabetes mellitus | | | | |
| Type 1 diabetes mellitus | 5013 (2.1) | 3028 (1.6) | 1985 (4.3) | 424 (6.8) |
| Type 2 diabetes mellitus | 36696 (15.5) | 22518 (11.8) | 14178 (30.6) | 2890 (46.4) |
| Chronic lower respiratory diseases | | | | |
| Bronchitis; not specified as acute or chronic | 11033 (4.7) | 8795 (4.6) | 2238 (4.8) | 362 (5.8) |
| Simple and mucopurulent chronic bronchitis | 988 (0.4) | 686 (0.4) | 302 (0.7) | 69 (1.1) |
| Unspecified chronic bronchitis | 1190 (0.5) | 734 (0.4) | 456 (1.0) | 81 (1.3) |
| Emphysema | 3454 (1.5) | 1947 (1.0) | 1507 (3.3) | 314 (5.0) |
| Other chronic obstructive pulmonary disease | 10564 (4.5) | 5714 (3.0) | 4850 (10.5) | 1030 (16.5) |
| Asthma | 25104 (10.6) | 19834 (10.4) | 5270 (11.4) | 755 (12.1) |
| Bronchiectasis | 1183 (0.5) | 719 (0.4) | 464 (1.0) | 89 (1.4) |

| | | | | |
|---|---|---|---|---|
| Nicotine dependence | 17105 (7.2) | 12639 (6.6) | 4466 (9.6) | 803 (12.9) |
| Substance misuse | 24870 (10.5) | 18173 ( 9.6) | 6697 (14.5) | 1316 (21.1) |
| Heart disease | | | | |
| Ischemic heart diseases | 21082 (8.9) | 11815 (6.2) | 9267 (20.0) | 2200 (35.3) |
| Other forms of heart disease | 42431 (18.0) | 26066 (13.7) | 16365 (35.3) | 3694 (59.3) |
| Chronic kidney diseases | | | | |
| Chronic kidney disease (CKD) | 15908 (6.7) | 8345 (4.4) | 7563 (16.3) | 1892 (30.4) |
| Hypertensive chronic kidney disease | 8943 (3.8) | 4460 (2.3) | 4483 (9.7) | 1247 (20.0) |
| Chronic liver disease | | | | |
| Alcoholic liver disease | 1091 (0.5) | 559 (0.3) | 532 (1.1) | 134 (2.2) |
| Hepatic failure; not elsewhere classified | 1409 (0.6) | 621 (0.3) | 788 (1.7) | 285 (4.6) |
| Chronic hepatitis; not elsewhere classified | 309 (0.1) | 200 (0.1) | 109 (0.2) | 29 (0.5) |
| Fibrosis and cirrhosis of liver | 2532 (1.1) | 1399 (0.7) | 1133 (2.4) | 248 (4.0) |
| Fatty (change of) liver; not elsewhere classified | 8190 (3.5) | 5882 (3.1) | 2308 (5.0) | 353 (5.7) |
| Chronic passive congestion of liver | 1330 (0.6) | 930 (0.5) | 400 (0.9) | 85 (1.4) |
| Portal hypertension | 1046 (0.4) | 540 (0.3) | 506 (1.1) | 120 (1.9) |
| Other specified diseases of liver | 5369 (2.3) | 3813 (2.0) | 1556 (3.4) | 293 (4.7) |
| Cerebral infarction | 5858 (2.5) | 3178 (1.7) | 2680 (5.8) | 925 (14.8) |
| Dementia | | | | |
| Vascular dementia | 1292 (0.5) | 648 (0.3) | 644 (1.4) | 332 (5.3) |
| Dementia in other diseases classified elsewhere | 2113 (0.9) | 1109 (0.6) | 1004 (2.2) | 551 (8.8) |
| Unspecified dementia | 4625 (2.0) | 2144 (1.1) | 2481 (5.4) | 1202 (19.3) |
| Alzheimer disease | 1652 (0.7) | 884 (0.5) | 768 (1.7) | 371 (6.0) |
| Frontotemporal dementia | 102 (0.04) | 57 (0.03) | 45 (0.1) | 23 (0.4) |
| Dementia with Lewy bodies | 139 (0.06) | 75 (0.04) | 64 (0.1) | 45 (0.7) |
| Neoplasms | | | | |
| Neoplasms (any) | 45255 (19.1) | 34362 (18.1) | 10893 (23.5) | 1793 (28.8) |
| Malignant neoplasms of lymphoid; hematopoietic and related tissue | 2655 (1.12) | 1598 (0.8) | 1057 (2.3) | 162 (2.6) |
| Organ transplant | | | | |
| Renal Transplantation Procedures | 741 (0.3) | 351 (0.2) | 390 (0.8) | 46 (0.7) |
| Liver Transplantation Procedures | 145 (0.1) | 73 (0.04) | 72 (0.2) | 11 (0.2) |
| Psoriasis | 2528 (1.1) | 1968 (1.0) | 560 (1.2) | 83 (1.3) |
| Rheumatoid arthritis | | | | |
| Rheumatoid arthritis with rheumatoid factor | 972 (0.4) | 732 (0.4) | 240 (0.5) | 40 (0.6) |
| Other rheumatoid arthritis | 3328 (1.4) | 2299 (1.2) | 1029 (2.2) | 175 (2.8) |
| Systemic lupus erythematosus (SLE) | 1417 (0.6) | 1046 (0.6) | 371 (0.8) | 51 (0.8) |
| Disorders involving the immune mechanism | 5215 (2.2) | 3437 (1.8) | 1778 (3.8) | 269 (4.3) |
| OUTCOMES | | | | |
| Any | 33.62 (33.17-34.07) | 31.74 (31.22-32.27) | 38.73 (37.87-39.60) | 62.34 (60.14-64.55) |
| Any (excluding dementia) | 32.59 (32.16-33.03) | 31.18 (30.67-31.70) | 36.32 (35.50-37.15) | 55.41 (53.28-57.57) |

**Table S2** – Incidence of neurologic and psychiatric outcomes within 6 months after a diagnosis of COVID-19 separately estimated by gender, race and age group

| | **Female** | **Male** | **Asian** | **Black** | **White** | **Over 65** | **Under 65** |
|---|---|---|---|---|---|---|---|
| | % (95% CI) | % (95% CI) | % (95% CI) | % (95% CI) | % (95% CI) | % (95% CI) | % (95% CI) |
| Intracranial haemorrhage (any) | 0.40 (0.33-0.49) | 0.81 (0.68-0.97) | 1.09 (0.58-2.04) | 0.55 (0.43-0.70) | 0.56 (0.47-0.68) | 1.21 (1.02-1.44) | 0.35 (0.30-0.42) |
| Intracranial haemorrhage (first) | 0.18 (0.13-0.25) | 0.42 (0.33-0.54) | 0.62 (0.23-1.67) | 0.24 (0.16-0.35) | 0.28 (0.21-0.37) | 0.62 (0.48-0.81) | 0.16 (0.12-0.21) |
| Ischaemic stroke (any) | 1.73 (1.58-1.90) | 2.51 (2.29-2.75) | 2.01 (1.35-2.99) | 2.70 (2.42-3.00) | 1.82 (1.66-2.00) | 4.75 (4.38-5.14) | 1.15 (1.03-1.27) |
| Ischaemic stroke (first) | 0.64 (0.53-0.76) | 0.90 (0.76-1.06) | 1.11 (0.62-1.97) | 0.96 (0.78-1.18) | 0.59 (0.49-0.71) | 1.65 (1.40-1.94) | 0.45 (0.38-0.54) |
| Parkinsonism | 0.045 (0.029-0.069) | 0.19 (0.13-0.28) | 0.09 (0.027-0.30) | 0.078 (0.033-0.18) | 0.11 (0.077-0.16) | 0.35 (0.25-0.48) | 0.022 (0.011-0.046) |
| Guillain-Barre syndrome | 0.073 (0.049-0.11) | 0.11 (0.074-0.15) | 0.15 (0.041-0.55) | 0.033 (0.016-0.07) | 0.10 (0.074-0.15) | 0.13 (0.078-0.21) | 0.07 (0.051-0.095) |
| Nerve/nerve root/plexus disorders | 3.15 (2.92-3.39) | 2.46 (2.21-2.73) | 2.29 (1.60-3.28) | 2.99 (2.66-3.35) | 2.92 (2.69-3.17) | 3.35 (3.00-3.73) | 2.65 (2.46-2.85) |
| Myoneural junction/muscle disease | 0.33 (0.27-0.41) | 0.63 (0.52-0.77) | 0.38 (0.20-0.74) | 0.40 (0.29-0.54) | 0.42 (0.34-0.51) | 0.61 (0.49-0.76) | 0.39 (0.33-0.48) |
| Encephalitis | 0.087 (0.058-0.13) | 0.13 (0.085-0.19) | 0.28 (0.099-0.77) | 0.13 (0.073-0.24) | 0.094 (0.066-0.13) | 0.15 (0.095-0.24) | 0.093 (0.064-0.13) |
| Dementia | 0.69 (0.59-0.82) | 0.65 (0.54-0.78) | 0.62 (0.32-1.19) | 0.67 (0.53-0.85) | 0.75 (0.63-0.88) | 2.66 (2.35-3.02) | 0.10 (0.069-0.14) |
| Mood/Anxiety/Psychotic disorder (any) | 28.00 (27.43-28.57) | 18.38 (17.78-18.99) | 15.71 (13.81-17.85) | 21.05 (20.29-21.84) | 27.47 (26.87-28.09) | 23.42 (22.65-24.22) | 23.85 (23.36-24.35) |
| Mood/Anxiety/Psychotic disorder (first) | 10.08 (9.55-10.62) | 6.91 (6.44-7.42) | 7.31 (5.74-9.29) | 7.95 (7.30-8.65) | 9.43 (8.89-10.01) | 7.68 (7.07-8.34) | 8.75 (8.33-9.20) |
| Mood disorder (any) | 16.20 (15.74-16.67) | 9.92 (9.46-10.40) | 8.45 (6.92-10.31) | 11.66 (11.07-12.29) | 15.86 (15.37-16.36) | 14.52 (13.89-15.18) | 13.08 (12.70-13.48) |
| Mood disorder (first) | 5.07 (4.72-5.43) | 3.26 (2.93-3.64) | 3.38 (2.23-5.10) | 3.71 (3.27-4.20) | 4.75 (4.40-5.14) | 4.07 (3.63-4.57) | 4.24 (3.96-4.54) |
| Anxiety disorder (any) | 20.94 (20.44-21.46) | 12.50 (11.99-13.03) | 10.87 (9.25-12.75) | 14.90 (14.24-15.60) | 20.08 (19.54-20.62) | 14.21 (13.58-14.87) | 18.26 (17.82-18.70) |
| Anxiety disorder (first) | 8.52 (8.08-8.99) | 5.33 (4.94-5.74) | 6.30 (4.82-8.21) | 6.59 (6.05-7.18) | 7.66 (7.21-8.15) | 5.48 (4.99-6.02) | 7.50 (7.14-7.88) |
| Psychotic disorder (any) | 1.22 (1.10-1.37) | 1.81 (1.62-2.03) | 1.05 (0.63-1.76) | 1.89 (1.67-2.15) | 1.42 (1.27-1.60) | 2.27 (2.01-2.58) | 1.16 (1.05-1.28) |
| Psychotic disorder (first) | 0.38 (0.30-0.48) | 0.52 (0.41-0.65) | 0.30 (0.13-0.67) | 0.52 (0.40-0.69) | 0.45 (0.36-0.57) | 0.82 (0.65-1.03) | 0.30 (0.24-0.38) |
| Substance misuse (any) | 5.45 (5.18-5.74) | 8.48 (8.07-8.91) | 1.86 (1.36-2.54) | 8.29 (7.79-8.82) | 6.68 (6.37-7.00) | 5.32 (4.91-5.77) | 6.88 (6.61-7.16) |
| Substance misuse (first) | 1.42 (1.26-1.61) | 2.59 (2.32-2.89) | 0.65 (0.37-1.12) | 2.22 (1.89-2.60) | 1.84 (1.65-2.04) | 1.46 (1.22-1.73) | 1.95 (1.78-2.14) |
| Insomnia (any) | 5.80 (5.50-6.12) | 5.07 (4.73-5.44) | 4.51 (3.31-6.12) | 5.03 (4.62-5.47) | 6.03 (5.71-6.37) | 6.87 (6.40-7.38) | 4.95 (4.69-5.21) |
| Insomnia (first) | 2.73 (2.49-2.99) | 2.28 (2.05-2.53) | 2.67 (1.68-4.25) | 2.46 (2.16-2.81) | 2.60 (2.37-2.86) | 2.87 (2.53-3.26) | 2.39 (2.20-2.60) |

**Table S3** – Characteristics of the COVID-19 and influenza cohorts after propensity-score matching. SMD=Standardized Mean Difference.

| | **COVID-19** | **Influenza** | **SMD** |
|---|---|---|---|
| Number | 105579 | 105579 | - |
| DEMOGRAPHICS | | | |
| Age; mean (SD); y | 39.7 (18.4) | 38.6 (19.7) | 0.06 |
| Sex; n (%) | | | |
| Female | 61831 (58.6) | 60828 (57.6) | 0.02 |
| Male | 43709 (41.4) | 44707 (42.3) | 0.02 |
| Other | 39 (0.04) | 44 (0.04) | 0.002 |
| Race; n (%) | | | |
| White | 69730 (66.0) | 69294 (65.6) | 0.009 |
| Black or African American | 19175 (18.2) | 18466 (17.5) | 0.02 |
| Asian | 3724 (3.5) | 3488 (3.3) | 0.01 |
| American Indian or Alaska Native | 442 (0.4) | 431 (0.4) | 0.002 |
| Native Hawaiian or Other Pacific Islander | 214 (0.2) | 219 (0.2) | 0.001 |
| Unknown | 12294 (11.6) | 13681 (13.0) | 0.04 |
| Ethnicity; n (%) | | | |
| Hispanic or Latino | 8841 (8.4) | 8932 (8.5) | 0.003 |
| Not Hispanic of Latino | 72389 (68.6) | 71409 (67.6) | 0.02 |
| Unknown | 24349 (23.1) | 25238 (23.9) | 0.02 |
| Problems related to housing and economic circumstances; n (%) | 937 (0.9) | 875 (0.8) | 0.006 |
| COMORBIDITIES; n (%) | | | |
| Overweight and obesity | 19293 (18.3) | 18078 (17.1) | 0.03 |
| Hypertensive disease | 28338 (26.8) | 26182 (24.8) | 0.05 |
| Diabetes mellitus | | | |
| Type 1 diabetes mellitus | 1995 (1.9) | 1894 (1.8) | 0.007 |
| Type 2 diabetes mellitus | 12133 (11.5) | 11300 (10.7) | 0.03 |
| Chronic lower respiratory diseases | | | |
| Simple and mucopurulent chronic bronchitis | 624 (0.6) | 585 (0.6) | 0.005 |
| Unspecified chronic bronchitis | 659 (0.6) | 643 (0.6) | 0.002 |
| Emphysema | 1779 (1.7) | 1706 (1.6) | 0.005 |
| Other chronic obstructive pulmonary disease | 5699 (5.4) | 5498 (5.2) | 0.008 |
| Asthma | 16643 (15.8) | 15963 (15.1) | 0.02 |
| Bronchiectasis | 672 (0.6) | 656 (0.6) | 0.002 |
| Nicotine dependence | 12134 (11.5) | 11839 (11.2) | 0.009 |
| Substance misuse | 15716 (14.9) | 15182 (14.4) | 0.01 |
| Heart disease | | | |
| Ischemic heart diseases | 7719 (7.3) | 7340 (7.0) | 0.01 |
| Other forms of heart disease | 16717 (15.8) | 15685 (14.9) | 0.03 |

| | | | |
|---|---|---|---|
| Chronic kidney diseases | | | |
| Chronic kidney disease (CKD) | 5471 (5.2) | 5090 (4.8) | 0.02 |
| Hypertensive chronic kidney disease | 3158 (3.0) | 2904 (2.8) | 0.01 |
| Chronic liver disease | | | |
| Alcoholic liver disease | 366 (0.3) | 335 (0.3) | 0.005 |
| Fibrosis and cirrhosis of liver | 907 (0.9) | 820 (0.8) | 0.009 |
| Chronic passive congestion of liver | 668 (0.6) | 635 (0.6) | 0.004 |
| Portal hypertension | 333 (0.3) | 309 (0.3) | 0.004 |
| Other specified diseases of liver | 2276 (2.2) | 2168 (2.1) | 0.007 |
| Cerebral infarction | 1780 (1.7) | 1676 (1.6) | 0.008 |
| Dementia | | | |
| Vascular dementia | 234 (0.2) | 196 (0.2) | 0.008 |
| Dementia in other diseases classified elsewhere | 382 (0.4) | 339 (0.3) | 0.007 |
| Unspecified dementia | 771 (0.7) | 714 (0.7) | 0.006 |
| Alzheimer disease | 287 (0.3) | 257 (0.2) | 0.006 |
| Frontotemporal dementia | 16 (0.01) | 14 (0.01) | 0.002 |
| Dementia with Lewy bodies | 22 (0.02) | 26 (0.03) | 0.003 |
| Neoplasms | | | |
| Neoplasms (any) | 20992 (19.9) | 19481 (18.5) | 0.04 |
| Organ transplant | | | |
| Renal Transplantation Procedures | 208 (0.2) | 179 (0.2) | 0.006 |
| Liver Transplantation Procedures | 51 (0.05) | 42 (0.04) | 0.004 |
| Psoriasis | 1359 (1.3) | 1305 (1.2) | 0.005 |
| Rheumatoid arthritis | | | |
| Rheumatoid arthritis with rheumatoid factor | 481 (0.5) | 474 (0.4) | 0.001 |
| Other rheumatoid arthritis | 1691 (1.6) | 1572 (1.5) | 0.009 |
| Systemic lupus erythematosus (SLE) | 707 (0.7) | 673 (0.6) | 0.004 |
| Disorders involving the immune mechanism | 2609 (2.5) | 2502 (2.4) | 0.007 |

**Table S4** – Characteristics of the COVID-19 and other respiratory tract infection (RTI) cohorts after propensity-score matching. SMD=Standardized Mean Difference.

| | COVID-19 | Other RTI | SMD |
|---|---|---|---|
| Number | 236038 | 236038 | - |
| DEMOGRAPHICS | | | |
| Age; mean (SD); y | 45.9 (19.7) | 46.0 (20.4) | 0.005 |
| Sex; n (%) | | | |
| Female | 131376 (55.7) | 132927 (56.3) | 0.01 |
| Male | 103760 (44.0) | 102170 (43.3) | 0.01 |
| Other | 902 (0.4) | 941 (0.4) | 0.003 |
| Race; n (%) | | | |
| White | 135106 (57.2) | 137562 (58.3) | 0.02 |
| Black or African American | 44394 (18.8) | 42208 (17.9) | 0.02 |
| Asian | 6972 (3.0) | 6696 (2.8) | 0.007 |
| American Indian or Alaska Native | 960 (0.4) | 880 (0.4) | 0.005 |
| Native Hawaiian or Other Pacific Islander | 741 (0.3) | 706 (0.3) | 0.003 |
| Unknown | 47865 (20.3) | 47986 (20.3) | 0.001 |
| Ethnicity; n (%) | | | |
| Hispanic or Latino | 37496 (15.9) | 34960 (14.8) | 0.03 |
| Not Hispanic of Latino | 134034 (56.8) | 135033 (57.2) | 0.009 |
| Unknown | 64508 (27.3) | 66045 (28.0) | 0.01 |
| Problems related to housing and economic circumstances; n (%) | 2438 (1.0) | 2201 (0.9) | 0.01 |
| COMORBIDITIES; n (%) | | | |
| Overweight and obesity | 42807 (18.1) | 40343 (17.1) | 0.03 |
| Hypertensive disease | 70868 (30.0) | 66780 (28.3) | 0.04 |
| Diabetes mellitus | | | |
| Type 1 diabetes mellitus | 4993 (2.1) | 4468 (1.9) | 0.02 |
| Type 2 diabetes mellitus | 36535 (15.5) | 33881 (14.4) | 0.03 |
| Chronic lower respiratory diseases | | | |
| Simple and mucopurulent chronic bronchitis | 988 (0.4) | 1038 (0.4) | 0.003 |
| Unspecified chronic bronchitis | 1190 (0.5) | 1198 (0.5) | 5.00E-04 |
| Emphysema | 3452 (1.5) | 3434 (1.5) | 6.00E-04 |
| Other chronic obstructive pulmonary disease | 10562 (4.5) | 10301 (4.4) | 0.005 |
| Asthma | 25100 (10.6) | 24044 (10.2) | 0.01 |
| Bronchiectasis | 1183 (0.5) | 1172 (0.5) | 7.00E-04 |
| Nicotine dependence | 17105 (7.2) | 17173 (7.3) | 0.001 |
| Substance misuse | 24862 (10.5) | 24456 (10.4) | 0.006 |
| Heart disease | | | |
| Ischemic heart diseases | 21021 (8.9) | 19453 (8.2) | 0.02 |
| Other forms of heart disease | 42331 (17.9) | 39725 (16.8) | 0.03 |

| | | | |
|---|---|---|---|
| Chronic kidney diseases | | | |
| Chronic kidney disease (CKD) | 15837 (6.7) | 14406 (6.1) | 0.02 |
| Hypertensive chronic kidney disease | 8885 (3.8) | 8116 (3.4) | 0.02 |
| Chronic liver disease | | | |
| Alcoholic liver disease | 1089 (0.5) | 1018 (0.4) | 0.005 |
| Fibrosis and cirrhosis of liver | 2524 (1.1) | 2373 (1.0) | 0.006 |
| Chronic passive congestion of liver | 1329 (0.6) | 1217 (0.5) | 0.006 |
| Portal hypertension | 1042 (0.4) | 995 (0.4) | 0.003 |
| Other specified diseases of liver | 5363 (2.3) | 4994 (2.1) | 0.01 |
| Cerebral infarction | 5826 (2.5) | 5337 (2.3) | 0.01 |
| Dementia | | | |
| Vascular dementia | 1264 (0.5) | 1206 (0.5) | 0.003 |
| Dementia in other diseases classified elsewhere | 2077 (0.9) | 1948 (0.8) | 0.006 |
| Unspecified dementia | 4557 (1.9) | 4323 (1.8) | 0.007 |
| Alzheimer disease | 1629 (0.7) | 1544 (0.7) | 0.004 |
| Frontotemporal dementia | 100 (0.04) | 96 (0.04) | 8.00E-04 |
| Dementia with Lewy bodies | 137 (0.06) | 131 (0.06) | 0.001 |
| Neoplasms | | | |
| Neoplasms (any) | 45215 (19.2) | 43826 (18.6) | 0.02 |
| Organ transplant | | | |
| Renal Transplantation Procedures | 727 (0.3) | 640 (0.3) | 0.007 |
| Liver Transplantation Procedures | 143 (0.06) | 154 (0.07) | 0.002 |
| Psoriasis | 2528 (1.1) | 2366 (1.0) | 0.007 |
| Rheumatoid arthritis | | | |
| Rheumatoid arthritis with rheumatoid factor | 972 (0.4) | 847 (0.4) | 0.009 |
| Other rheumatoid arthritis | 3327 (1.4) | 3114 (1.3) | 0.008 |
| Systemic lupus erythematosus (SLE) | 1416 (0.6) | 1343 (0.6) | 0.004 |
| Disorders involving the immune mechanism | 5198 (2.2) | 4770 (2.0) | 0.01 |

**Table S5** – Hazard ratios and log-rank p-values estimated for the comparison between the COVID-19 cohort and the other two matched cohorts for subcategories of the main outcomes presented in Table 2 of the main manuscript. The corresponding ICD-10 codes are displayed in brackets.

| | **COVID-19 vs. Influenza** | | **COVID-19 vs. Other RTI** | |
|---|---|---|---|---|
| | HR (95% CI) | P-value | HR (95% CI) | P-value |
| Alcohol-related disorder (F10) | 1.71 (1.54-1.90) | <0.0001 | 1.30 (1.20-1.40) | <0.0001 |
| Opioid-related disorder (F11) | 1.53 (1.32-1.76) | <0.0001 | 1.16 (1.05-1.29) | 0.005 |
| Cannabis-related disorder (F12) | 1.33 (1.15-1.54) | 0.00012 | 1.09 (0.97-1.23) | 0.13 |
| Nicotine dependence (F17) | 1.14 (1.08-1.20) | <0.0001 | 0.98 (0.94-1.03) | 0.49 |
| Other substance misuse (F13–F16, F18, F19) | 1.55 (1.38-1.75) | <0.0001 | 1.15 (1.05-1.25) | 0.0021 |
| Cranial nerve disorder (G50-G53) | 1.36 (1.09-1.71) | 0.005 | 1.08 (0.93-1.24) | 0.3 |
| Nerve root and plexus disorder (G54-G55) | 1.76 (1.22-2.53) | 0.0017 | 1.53 (1.20-1.94) | 0.00039 |
| Mononeuropathies (G56-G59) | 1.68 (1.50-1.88) | <0.0001 | 1.33 (1.23-1.44) | <0.0001 |
| Myastenia gravis and other myoneural disorders (G70) | 3.88 (2.11-7.13) | <0.0001 | 2.19 (1.50-3.22) | <0.0001 |
| Myastenia gravis | 1.56 (0.55-4.46) | 0.67 | 0.96 (0.43-2.15) | 0.77 |
| Myoneural disorder; unspecified | 5.92 (2.99-11.75) | <0.0001 | 3.48 (2.16-5.61) | <0.0001 |
| Primary disorders of muscles (G71) | 0.43 (0.038-4.88) | 0.88 | 1.60 (0.53-4.82) | 0.34 |
| Other and unspecified myopathies (G72) | 6.02 (3.77-9.62) | <0.0001 | 4.31 (3.36-5.52) | <0.0001 |
| Specified myopathies (G72.0-G72.4) | 1.13 (0.35-3.60) | 0.61 | 1.29 (0.69-2.41) | 0.28 |
| Critical illness myopathy and other specified myopathies (G72.8) | 8.67 (4.75-15.81) | <0.0001 | 6.15 (4.50-8.40) | <0.0001 |
| Myopathy; unspecified (G72.9) | 3.09 (1.34-7.12) | 0.0051 | 3.00 (1.79-5.01) | <0.0001 |
| Myoneural junction/muscle disorder in diseases classified elsewhere (G73) | 4.4e-09 (0.00-Inf) | 0.39 | 1.24 (0.31-4.96) | 0.37 |
| Nontraumatic subarachnoid hemorrhage (I60) | 2.19 (1.37-3.50) | 0.00054 | 1.00 (0.79-1.27) | 0.92 |
| Nontraumatic intracerebral hemorrhage (I61) | 2.17 (1.51-3.11) | <0.0001 | 1.11 (0.91-1.34) | 0.28 |
| Other and unspecified nontraumatic intracranial hemorrhage (I62) | 3.01 (1.99-4.56) | <0.0001 | 1.32 (1.06-1.64) | 0.009 |

**Table S6** – Hazard ratios and log-rank p-values estimated for the comparison between the COVID-19 cohort and the additional four matched cohorts for the main neurologic and psychiatric outcomes presented in Table 2 of the main manuscript. PE=Pulmonary embolism

| | **Skin infection** (n=176079) | | **Urolithiasis** (n=125297) | | **Fracture** (n=162759) | | **PE** (n=51842) | |
|---|---|---|---|---|---|---|---|---|
| | HR (95% CI) | P-value | HR (95% CI) | P-value | HR (95% CI) | P-value | HR (95% CI) | P-value |
| Intracranial haemorrhage (any) | 1.63 (1.39-1.91) | <0.0001 | 1.43 (1.21-1.70) | <0.0001 | 0.45 (0.40-0.51) | <0.0001 | 0.60 (0.51-0.70) | <0.0001 |
| Intracranial haemorrhage (first) | 1.62 (1.28-2.05) | <0.0001 | 1.92 (1.47-2.53) | <0.0001 | 0.44 (0.36-0.53) | <0.0001 | 0.72 (0.55-0.94) | 0.017 |
| Ischaemic stroke (any) | 1.41 (1.30-1.52) | <0.0001 | 1.44 (1.32-1.56) | <0.0001 | 1.41 (1.31-1.52) | <0.0001 | 0.93 (0.86-1.01) | 0.1 |
| Ischaemic stroke (first) | 1.55 (1.35-1.78) | <0.0001 | 2.18 (1.84-2.58) | <0.0001 | 1.40 (1.22-1.60) | <0.0001 | 0.88 (0.74-1.05) | 0.16 |
| Parkinsonism | 1.28 (0.89-1.84) | 0.16 | 1.47 (1.01-2.13) | 0.036 | 0.78 (0.57-1.07) | 0.2 | 1.57 (0.99-2.49) | 0.039 |
| Guillain-Barre syndrome | 2.57 (1.64-4.03) | <0.0001 | 2.65 (1.62-4.33) | <0.0001 | 1.96 (1.32-2.91) | 0.00077 | 1.47 (0.91-2.37) | 0.11 |
| Nerve/nerve root/plexus disorders | 0.92 (0.86-0.98) | 0.012 | 1.19 (1.10-1.28) | <0.0001 | 0.61 (0.57-0.65) | <0.0001 | 1.10 (1.01-1.21) | 0.034 |
| Myoneural junction/muscle disease | 4.49 (3.51-5.74) | <0.0001 | 3.93 (3.04-5.09) | <0.0001 | 4.32 (3.37-5.54) | <0.0001 | 2.39 (1.82-3.15) | <0.0001 |
| Encephalitis | 1.50 (1.06-2.11) | 0.02 | 1.36 (0.92-2.01) | 0.094 | 1.69 (1.17-2.44) | 0.0041 | 0.87 (0.59-1.28) | 0.53 |
| Dementia | 1.48 (1.29-1.70) | <0.0001 | 3.38 (2.82-4.06) | <0.0001 | 1.06 (0.93-1.20) | 0.36 | 2.34 (1.95-2.82) | <0.0001 |
| Mood/Anxiety/Psychotic disorder (any) | 1.14 (1.12-1.17) | <0.0001 | 1.24 (1.21-1.27) | <0.0001 | 1.25 (1.22-1.28) | <0.0001 | 1.09 (1.06-1.12) | <0.0001 |
| Mood/Anxiety/Psychotic disorder (first) | 1.31 (1.25-1.38) | <0.0001 | 1.59 (1.50-1.69) | <0.0001 | 1.41 (1.34-1.48) | <0.0001 | 1.18 (1.08-1.28) | 0.00016 |
| Mood disorder (any) | 1.07 (1.04-1.10) | <0.0001 | 1.14 (1.10-1.17) | <0.0001 | 1.11 (1.07-1.14) | <0.0001 | 1.03 (0.99-1.08) | 0.089 |
| Mood disorder (first) | 1.09 (1.02-1.17) | 0.0068 | 1.28 (1.18-1.38) | <0.0001 | 1.17 (1.09-1.25) | <0.0001 | 1.04 (0.93-1.15) | 0.47 |
| Anxiety disorder (any) | 1.18 (1.15-1.21) | <0.0001 | 1.22 (1.19-1.26) | <0.0001 | 1.33 (1.30-1.37) | <0.0001 | 1.08 (1.04-1.12) | <0.0001 |
| Anxiety disorder (first) | 1.38 (1.31-1.45) | <0.0001 | 1.57 (1.47-1.67) | <0.0001 | 1.53 (1.45-1.62) | <0.0001 | 1.18 (1.08-1.29) | 0.00021 |
| Psychotic disorder (any) | 1.25 (1.15-1.36) | <0.0001 | 2.17 (1.93-2.44) | <0.0001 | 1.49 (1.36-1.63) | <0.0001 | 1.45 (1.28-1.64) | <0.0001 |
| Psychotic disorder (first) | 1.13 (0.95-1.35) | 0.14 | 2.43 (1.91-3.10) | <0.0001 | 1.30 (1.08-1.56) | 0.004 | 1.79 (1.34-2.38) | <0.0001 |
| Substance misuse (any) | 0.76 (0.74-0.79) | <0.0001 | 0.99 (0.95-1.04) | 0.79 | 0.85 (0.82-0.88) | <0.0001 | 0.85 (0.81-0.89) | <0.0001 |
| Substance misuse (first) | 0.53 (0.49-0.57) | <0.0001 | 0.76 (0.69-0.84) | <0.0001 | 0.52 (0.48-0.56) | <0.0001 | 0.65 (0.56-0.76) | <0.0001 |
| Insomnia (any) | 1.23 (1.18-1.30) | <0.0001 | 1.35 (1.28-1.43) | <0.0001 | 1.33 (1.26-1.40) | <0.0001 | 1.10 (1.04-1.18) | 0.0025 |
| Insomnia (first) | 1.49 (1.38-1.62) | <0.0001 | 1.68 (1.53-1.85) | <0.0001 | 1.46 (1.34-1.59) | <0.0001 | 1.06 (0.94-1.19) | 0.32 |

**Table S7** – P-values for the test of proportional hazards (obtained using the generalized Schoenfeld test) for the two primary control cohorts. A value lower than 0.05 indicates evidence for non-proportional hazards. RTI=Respiratory tract infections.

| | COVID-19 vs Influenza | COVID-19 vs Other RTI |
|---|---|---|
| Intracranial haemorrhage (any) | 0.11 | 0.012 |
| Intracranial haemorrhage (first) | 0.096 | 0.14 |
| Ischaemic stroke (any) | 0.24 | 0.032 |
| Ischaemic stroke (first) | 0.65 | 0.12 |
| Parkinsonism | 0.97 | 0.97 |
| Guillain-Barre syndrome | 0.076 | 0.11 |
| Nerve/nerve root/plexus disorders | 0.79 | 0.13 |
| Myoneural junction/muscle disease | 0.86 | 0.55 |
| Encephalitis | 0.62 | 0.28 |
| Dementia | 0.2 | 0.21 |
| Mood/Anxiety/Psychotic disorder (any) | <0.0001 | <0.0001 |
| Mood/Anxiety/Psychotic disorder (first) | 0.012 | 0.00052 |
| Mood disorder (any) | 0.0024 | <0.0001 |
| Mood disorder (first) | 0.48 | 0.89 |
| Anxiety disorder (any) | <0.0001 | <0.0001 |
| Anxiety disorder (first) | 0.0033 | <0.0001 |
| Psychotic disorder (any) | 0.12 | 0.78 |
| Psychotic disorder (first) | 0.87 | 0.036 |
| Substance misuse (any) | 0.92 | 0.56 |
| Substance misuse (first) | 0.4 | 0.1 |
| Insomnia (any) | 0.54 | 0.1 |
| Insomnia (first) | 0.039 | 0.059 |

**Table S8** – P-values for the test of proportional hazards (obtained using the generalized Schoenfeld test) for the other four control cohorts. A value lower than 0.05 indicates evidence for non-proportional hazards. RTI=Respiratory tract infections.

| | COVID-19 vs Skin Infection | COVID-19 vs Urolithiasis | COVID-19 vs Fracture | COVID-19 vs PE |
|---|---|---|---|---|
| Intracranial haemorrhage (any) | 0.9 | 0.059 | <0.0001 | 0.0017 |
| Intracranial haemorrhage (first) | 0.47 | 0.95 | <0.0001 | 0.011 |
| Ischaemic stroke (any) | 0.011 | 0.21 | 0.19 | <0.0001 |
| Ischaemic stroke (first) | 0.52 | 0.38 | 0.35 | 0.003 |
| Parkinsonism | 0.072 | 0.56 | 0.45 | 0.81 |
| Guillain-Barre syndrome | 0.023 | 0.71 | 0.0045 | 0.23 |
| Nerve/nerve root/plexus disorders | <0.0001 | <0.0001 | <0.0001 | <0.0001 |
| Myoneural junction/muscle disease | 0.55 | 1 | 0.19 | 0.83 |
| Encephalitis | 0.38 | 0.96 | 0.93 | 0.67 |
| Dementia | 0.00027 | 0.014 | 0.4 | 0.16 |
| Mood/Anxiety/Psychotic disorder (any) | 0.0016 | 0.45 | <0.0001 | <0.0001 |
| Mood/Anxiety/Psychotic disorder (first) | 0.57 | 0.045 | <0.0001 | 0.0048 |
| Mood disorder (any) | 0.44 | 0.44 | <0.0001 | 0.0043 |
| Mood disorder (first) | 0.0089 | 0.98 | <0.0001 | 0.00075 |
| Anxiety disorder (any) | 0.00011 | 0.6 | <0.0001 | <0.0001 |
| Anxiety disorder (first) | 0.0082 | 0.012 | 0.00055 | 0.037 |
| Psychotic disorder (any) | 0.025 | 0.00066 | 0.25 | 0.76 |
| Psychotic disorder (first) | 0.58 | 0.086 | 0.18 | 0.12 |
| Substance misuse (any) | <0.0001 | 0.58 | <0.0001 | <0.0001 |
| Substance misuse (first) | 0.0039 | 0.32 | <0.0001 | 0.0081 |
| Insomnia (any) | 0.49 | 0.33 | 0.0017 | 0.00034 |
| Insomnia (first) | 0.0059 | 0.025 | 0.0081 | 0.15 |

**Table S9** – Characteristics of the cohort of patients with COVID-19 not requiring hospitalization and the cohort of patients with influenza not requiring hospitalization, after propensity-score matching. SMD=Standardized Mean Difference.

| | **COVID-19 (non-hospitalized)** | **Influenza (non-hospitalized)** | **SMD** |
|---|---|---|---|
| Number | 96803 | 96803 | - |
| DEMOGRAPHICS | | | |
| Age; mean (SD); y | 38.6 (17.8) | 37.4 (19.0) | 0.07 |
| Sex; n (%) | | | |
| Female | 57328 (59.2) | 56090 (57.9) | 0.03 |
| Male | 39438 (40.7) | 40670 (42.0) | 0.03 |
| Other | 37 (0.04) | 43 (0.04) | 0.003 |
| Race; n (%) | | | |
| White | 63831 (65.9) | 63465 (65.6) | 0.008 |
| Black or African American | 17334 (17.9) | 16553 (17.1) | 0.02 |
| Asian | 3478 (3.6) | 3239 (3.3) | 0.01 |
| American Indian or Alaska Native | 404 (0.4) | 400 (0.4) | 6.00E-04 |
| Native Hawaiian or Other Pacific Islander | 274 (0.3) | 229 (0.2) | 0.009 |
| Unknown | 11482 (11.9) | 12917 (13.3) | 0.04 |
| Ethnicity; n (%) | | | |
| Hispanic or Latino | 8540 (8.8) | 8474 (8.8) | 0.002 |
| Not Hispanic of Latino | 65640 (67.8) | 64819 (67.0) | 0.02 |
| Unknown | 22623 (23.4) | 23510 (24.3) | 0.02 |
| Problems related to housing and economic circumstances; n (%) | 648 (0.7) | 595 (0.6) | 0.007 |
| COMORBIDITIES; n (%) | | | |
| Overweight and obesity | 16599 (17.1) | 15630 (16.1) | 0.03 |
| Hypertensive disease | 23247 (24.0) | 21322 (22.0) | 0.05 |
| Diabetes mellitus | | | |
| Type 1 diabetes mellitus | 1514 (1.6) | 1414 (1.5) | 0.008 |
| Type 2 diabetes mellitus | 9444 ( 9.8) | 8752 (9.0) | 0.02 |
| Chronic lower respiratory diseases | | | |
| Simple and mucopurulent chronic bronchitis | 464 (0.5) | 454 (0.5) | 0.002 |
| Unspecified chronic bronchitis | 458 (0.5) | 449 (0.5) | 0.001 |
| Emphysema | 1107 (1.1) | 1054 (1.1) | 0.005 |
| Other chronic obstructive pulmonary disease | 3458 (3.6) | 3372 (3.5) | 0.005 |
| Asthma | 14453 (14.9) | 14020 (14.5) | 0.01 |
| Bronchiectasis | 450 (0.5) | 438 (0.5) | 0.002 |
| Nicotine dependence | 9847 (10.2) | 9693 (10.0) | 0.005 |
| Substance misuse | 12840 (13.3) | 12492 (12.9) | 0.01 |
| Heart disease | | | |
| Ischemic heart diseases | 5456 (5.6) | 4982 (5.1) | 0.02 |

| | | | |
|---|---|---|---|
| Other forms of heart disease | 12647 (13.1) | 11768 (12.2) | 0.03 |
| Chronic kidney diseases | | | |
| Chronic kidney disease (CKD) | 3608 (3.7) | 3374 (3.5) | 0.01 |
| Hypertensive chronic kidney disease | 1966 (2.0) | 1820 (1.9) | 0.01 |
| Chronic liver disease | | | |
| Alcoholic liver disease | 255 (0.3) | 224 (0.2) | 0.006 |
| Fibrosis and cirrhosis of liver | 659 (0.7) | 578 (0.6) | 0.01 |
| Chronic passive congestion of liver | 516 (0.5) | 496 (0.5) | 0.003 |
| Portal hypertension | 246 (0.3) | 203 (0.2) | 0.009 |
| Other specified diseases of liver | 1862 (1.9) | 1754 (1.8) | 0.008 |
| Cerebral infarction | 1283 (1.3) | 1135 (1.2) | 0.01 |
| Dementia | | | |
| Vascular dementia | 142 (0.1) | 129 (0.1) | 0.004 |
| Dementia in other diseases classified elsewhere | 242 (0.2) | 213 (0.2) | 0.006 |
| Unspecified dementia | 446 (0.5) | 388 (0.4) | 0.009 |
| Alzheimer disease | 173 (0.2) | 156 (0.2) | 0.004 |
| Frontotemporal dementia | 17 (0.02) | 10 (0.01) | 0.006 |
| Dementia with Lewy bodies | 27 (0.03) | 18 (0.02) | 0.006 |
| Neoplasms | | | |
| Neoplasms (any) | 18338 (18.9) | 17048 (17.6) | 0.03 |
| Organ transplant | | | |
| Renal Transplantation Procedures | 100 (0.1) | 106 (0.1) | 0.002 |
| Liver Transplantation Procedures | 29 (0.03) | 22 (0.02) | 0.004 |
| Psoriasis | 1193 (1.2) | 1171 (1.2) | 0.002 |
| Rheumatoid arthritis | | | |
| Rheumatoid arthritis with rheumatoid factor | 392 (0.4) | 384 (0.4) | 0.001 |
| Other rheumatoid arthritis | 1283 (1.3) | 1249 (1.3) | 0.003 |
| Systemic lupus erythematosus (SLE) | 606 (0.6) | 564 (0.6) | 0.006 |
| Disorders involving the immune mechanism | 1987 (2.1) | 1887 (1.9) | 0.007 |

**Table S10** – Characteristics of the cohort of patients with COVID-19 not requiring hospitalization and the cohort of patients with other respiratory tract infections (RTI) not requiring hospitalization, after propensity-score matching. SMD=Standardized Mean Difference.

| | **COVID-19 (non-hospitalized)** | **Other RTI (non-hospitalized)** | **SMD** |
|---|---|---|---|
| Number | 183731 | 183731 | - |
| DEMOGRAPHICS | | | |
| Age; mean (SD); y | 43.3 (19.0) | 43.3 (19.6) | 7.00E-04 |
| Sex; n (%) | | | |
| Female | 104084 (56.6) | 105084 (57.2) | 0.01 |
| Male | 78969 (43.0) | 77943 (42.4) | 0.01 |
| Other | 678 (0.4) | 704 (0.4) | 0.002 |
| Race; n (%) | | | |
| White | 105595 (57.5) | 107903 (58.7) | 0.03 |
| Black or African American | 32587 (17.7) | 30676 (16.7) | 0.03 |
| Asian | 5291 (2.9) | 5083 (2.8) | 0.007 |
| American Indian or Alaska Native | 702 (0.4) | 586 (0.3) | 0.01 |
| Native Hawaiian or Other Pacific Islander | 814 (0.4) | 792 (0.4) | 0.002 |
| Unknown | 38742 (21.1) | 38691 (21.1) | 7.00E-04 |
| Ethnicity; n (%) | | | |
| Hispanic or Latino | 27936 (15.2) | 25912 (14.1) | 0.03 |
| Not Hispanic of Latino | 102332 (55.7) | 103612 (56.4) | 0.01 |
| Unknown | 53463 (29.1) | 54207 (29.5) | 0.009 |
| Problems related to housing and economic circumstances; n (%) | 1371 (0.7) | 1232 (0.7) | 0.009 |
| COMORBIDITIES; n (%) | | | |
| Overweight and obesity | 29583 (16.1) | 27662 (15.1) | 0.03 |
| Hypertensive disease | 46219 (25.2) | 42968 (23.4) | 0.04 |
| Diabetes mellitus | | | |
| Type 1 diabetes mellitus | 2922 (1.6) | 2572 (1.4) | 0.02 |
| Type 2 diabetes mellitus | 21968 (12.0) | 19685 (10.7) | 0.04 |
| Chronic lower respiratory diseases | | | |
| Simple and mucopurulent chronic bronchitis | 653 (0.4) | 720 (0.4) | 0.006 |
| Unspecified chronic bronchitis | 711 (0.4) | 793 (0.4) | 0.007 |
| Emphysema | 1891 (1.0) | 1895 (1.0) | 2.00E-04 |
| Other chronic obstructive pulmonary disease | 5669 (3.1) | 5627 (3.1) | 0.001 |
| Asthma | 19226 (10.5) | 18273 ( 9.9) | 0.02 |
| Bronchiectasis | 691 (0.4) | 730 (0.4) | 0.003 |
| Nicotine dependence | 12391 (6.7) | 12345 (6.7) | 0.001 |
| Substance misuse | 17747 ( 9.7) | 17330 (9.4) | 0.008 |
| Heart disease | | | |
| Ischemic heart diseases | 11671 (6.4) | 10483 (5.7) | 0.03 |

| | | | |
|---|---|---|---|
| Other forms of heart disease | 25371 (13.8) | 23320 (12.7) | 0.03 |
| Chronic kidney diseases | | | |
| Chronic kidney disease (CKD) | 8205 (4.5) | 7221 (3.9) | 0.03 |
| Hypertensive chronic kidney disease | 4437 (2.4) | 3905 (2.1) | 0.02 |
| Chronic liver disease | | | |
| Alcoholic liver disease | 557 (0.3) | 499 (0.3) | 0.006 |
| Fibrosis and cirrhosis of liver | 1373 (0.7) | 1219 (0.7) | 0.01 |
| Chronic passive congestion of liver | 889 (0.5) | 841 (0.5) | 0.004 |
| Portal hypertension | 526 (0.3) | 468 (0.3) | 0.006 |
| Other specified diseases of liver | 3623 (2.0) | 3229 (1.8) | 0.02 |
| Cerebral infarction | 3093 (1.7) | 2686 (1.5) | 0.02 |
| Dementia | | | |
| Vascular dementia | 640 (0.3) | 609 (0.3) | 0.003 |
| Dementia in other diseases classified elsewhere | 1098 (0.6) | 1002 (0.5) | 0.007 |
| Unspecified dementia | 2157 (1.2) | 1997 (1.1) | 0.008 |
| Alzheimer disease | 867 (0.5) | 815 (0.4) | 0.004 |
| Frontotemporal dementia | 57 (0.03) | 40 (0.02) | 0.006 |
| Dementia with Lewy bodies | 79 (0.04) | 70 (0.04) | 0.002 |
| Neoplasms | | | |
| Neoplasms (any) | 33036 (18.0) | 31974 (17.4) | 0.02 |
| Organ transplant | | | |
| Renal Transplantation Procedures | 328 (0.2) | 283 (0.2) | 0.006 |
| Liver Transplantation Procedures | 66 (0.04) | 67 (0.04) | 3.00E-04 |
| Psoriasis | 1896 (1.0) | 1701 (0.9) | 0.01 |
| Rheumatoid arthritis | | | |
| Rheumatoid arthritis with rheumatoid factor | 704 (0.4) | 672 (0.4) | 0.003 |
| Other rheumatoid arthritis | 2220 (1.2) | 2075 (1.1) | 0.007 |
| Systemic lupus erythematosus (SLE) | 997 (0.5) | 876 (0.5) | 0.009 |
| Disorders involving the immune mechanism | 3321 (1.8) | 2974 (1.6) | 0.01 |

**Table S11** – Characteristics of the cohort of patients with COVID-19 requiring hospitalization and the cohort of patients with COVID-19 not requiring hospitalization, after propensity-score matching. SMD=Standardized Mean Difference.

| | COVID-19 with hospitalization | COVID-19 without hospitalization | SMD |
|---|---|---|---|
| Number | 44927 | 44927 | - |
| DEMOGRAPHICS | | | |
| Age; mean (SD); y | 56.4 (18.7) | 56.8 (18.2) | 0.02 |
| Sex; n (%) | | | |
| Female | 23161 (51.6) | 24064 (53.6) | 0.04 |
| Male | 21697 (48.3) | 20834 (46.4) | 0.04 |
| Other | 69 (0.2) | 29 (0.07) | 0.03 |
| Race; n (%) | | | |
| White | 24757 (55.1) | 25081 (55.8) | 0.01 |
| Black or African American | 10150 (22.6) | 10395 (23.1) | 0.01 |
| Asian | 1511 (3.4) | 1548 (3.4) | 0.005 |
| American Indian or Alaska Native | 243 (0.5) | 248 (0.6) | 0.002 |
| Native Hawaiian or Other Pacific Islander | 162 (0.4) | 144 (0.3) | 0.007 |
| Unknown | 8104 (18.0) | 7511 (16.7) | 0.03 |
| Ethnicity; n (%) | | | |
| Hispanic or Latino | 8338 (18.6) | 8779 (19.5) | 0.02 |
| Not Hispanic of Latino | 26296 (58.5) | 27081 (60.3) | 0.04 |
| Unknown | 10293 (22.9) | 9067 (20.2) | 0.07 |
| Problems related to housing and economic circumstances; n (%) | 941 (2.1) | 909 (2.0) | 0.005 |
| COMORBIDITIES; n (%) | | | |
| Overweight and obesity | 11981 (26.7) | 12624 (28.1) | 0.03 |
| Hypertensive disease | 22199 (49.4) | 22687 (50.5) | 0.02 |
| Diabetes mellitus | | | |
| Type 1 diabetes mellitus | 1779 (4.0) | 1774 (3.9) | 6.00E-04 |
| Type 2 diabetes mellitus | 13127 (29.2) | 13105 (29.2) | 0.001 |
| Chronic lower respiratory diseases | | | |
| Simple and mucopurulent chronic bronchitis | 292 (0.7) | 300 (0.7) | 0.002 |
| Unspecified chronic bronchitis | 420 (0.9) | 411 (0.9) | 0.002 |
| Emphysema | 1365 (3.0) | 1277 (2.8) | 0.01 |
| Other chronic obstructive pulmonary disease | 4300 ( 9.6) | 3958 (8.8) | 0.03 |
| Asthma | 5070 (11.3) | 5342 (11.9) | 0.02 |
| Bronchiectasis | 426 (0.9) | 419 (0.9) | 0.002 |
| Nicotine dependence | 4222 (9.4) | 4263 (9.5) | 0.003 |
| Substance misuse | 6301 (14.0) | 6453 (14.4) | 0.01 |
| Heart disease | | | |
| Ischemic heart diseases | 8382 (18.7) | 7973 (17.7) | 0.02 |

| | | | |
|---|---|---|---|
| Other forms of heart disease | 15111 (33.6) | 14913 (33.2) | 0.009 |
| Chronic kidney diseases | | | |
| Chronic kidney disease (CKD) | 6675 (14.9) | 6105 (13.6) | 0.04 |
| Hypertensive chronic kidney disease | 3935 (8.8) | 3498 (7.8) | 0.04 |
| Chronic liver disease | | | |
| Alcoholic liver disease | 453 (1.0) | 409 (0.9) | 0.01 |
| Fibrosis and cirrhosis of liver | 991 (2.2) | 900 (2.0) | 0.01 |
| Chronic passive congestion of liver | 373 (0.8) | 369 (0.8) | 0.001 |
| Portal hypertension | 439 (1.0) | 387 (0.9) | 0.01 |
| Other specified diseases of liver | 1464 (3.3) | 1479 (3.3) | 0.002 |
| Cerebral infarction | 2387 (5.3) | 2187 (4.9) | 0.02 |
| Dementia | | | |
| Vascular dementia | 559 (1.2) | 502 (1.1) | 0.01 |
| Dementia in other diseases classified elsewhere | 901 (2.0) | 853 (1.9) | 0.008 |
| Unspecified dementia | 2130 (4.7) | 1825 (4.1) | 0.03 |
| Alzheimer disease | 696 (1.5) | 664 (1.5) | 0.006 |
| Frontotemporal dementia | 44 (0.1) | 45 (0.1) | 7.00E-04 |
| Dementia with Lewy bodies | 55 (0.1) | 51 (0.1) | 0.003 |
| Neoplasms | | | |
| Neoplasms (any) | 10456 (23.3) | 10969 (24.4) | 0.03 |
| Organ transplant | | | |
| Renal Transplantation Procedures | 327 (0.7) | 292 (0.7) | 0.009 |
| Liver Transplantation Procedures | 63 (0.1) | 55 (0.1) | 0.005 |
| Psoriasis | 538 (1.2) | 524 (1.2) | 0.003 |
| Rheumatoid arthritis | | | |
| Rheumatoid arthritis with rheumatoid factor | 230 (0.5) | 244 (0.5) | 0.004 |
| Other rheumatoid arthritis | 960 (2.1) | 997 (2.2) | 0.006 |
| Systemic lupus erythematosus (SLE) | 359 (0.8) | 367 (0.8) | 0.002 |
| Disorders involving the immune mechanism | 1658 (3.7) | 1658 (3.7) | 0 |

**Table S12** – Characteristics of the cohort of patients with COVID-19 and encephalopathy and the cohort of patients with COVID-19 but without encephalopathy, after propensity-score matching. SMD=Standardized Mean Difference.

| | COVID-19 with encephalopathy | COVID-19 without encephalopathy | SMD |
|---|---|---|---|
| Number | 6221 | 6221 | - |
| DEMOGRAPHICS | | | |
| Age; mean (SD); y | 66.7 (17.0) | 67.6 (16.0) | 0.05 |
| Sex; n (%) | | | |
| Female | 2908 (46.7) | 2929 (47.1) | 0.007 |
| Male | 3300 (53.0) | 3288 (52.9) | 0.004 |
| Other | 13 (0.2) | 10 (0.2) | 0.01 |
| Race; n (%) | | | |
| White | 3330 (53.5) | 3437 (55.2) | 0.03 |
| Black or African American | 1549 (24.9) | 1528 (24.6) | 0.008 |
| Asian | 242 (3.9) | 235 (3.8) | 0.006 |
| American Indian or Alaska Native | 21 (0.3) | 22 (0.4) | 0.003 |
| Native Hawaiian or Other Pacific Islander | 11 (0.2) | 10 (0.2) | 0.004 |
| Unknown | 1068 (17.2) | 990 (15.9) | 0.03 |
| Ethnicity; n (%) | | | |
| Hispanic or Latino | 894 (14.4) | 928 (14.9) | 0.02 |
| Not Hispanic of Latino | 3867 (62.2) | 3875 (62.3) | 0.003 |
| Unknown | 1460 (23.5) | 1418 (22.8) | 0.02 |
| Problems related to housing and economic circumstances; n (%) | 215 (3.5) | 195 (3.1) | 0.02 |
| COMORBIDITIES; n (%) | | | |
| Overweight and obesity | 1832 (29.4) | 2002 (32.2) | 0.06 |
| Hypertensive disease | 4583 (73.7) | 4853 (78.0) | 0.1 |
| Diabetes mellitus | | | |
| Type 1 diabetes mellitus | 422 (6.8) | 439 (7.1) | 0.01 |
| Type 2 diabetes mellitus | 2884 (46.4) | 2986 (48.0) | 0.03 |
| Chronic lower respiratory diseases | | | |
| Simple and mucopurulent chronic bronchitis | 69 (1.1) | 84 (1.4) | 0.02 |
| Unspecified chronic bronchitis | 81 (1.3) | 105 (1.7) | 0.03 |
| Emphysema | 314 (5.0) | 305 (4.9) | 0.007 |
| Other chronic obstructive pulmonary disease | 1028 (16.5) | 1055 (17.0) | 0.01 |
| Asthma | 753 (12.1) | 815 (13.1) | 0.03 |
| Bronchiectasis | 89 (1.4) | 97 (1.6) | 0.01 |
| Nicotine dependence | 802 (12.9) | 821 (13.2) | 0.009 |
| Substance misuse | 1310 (21.1) | 1347 (21.7) | 0.01 |
| Heart disease | | | |

| | | | |
|---|---|---|---|
| Ischemic heart diseases | 2193 (35.3) | 2221 (35.7) | 0.009 |
| Other forms of heart disease | 3686 (59.3) | 3834 (61.6) | 0.05 |
| Chronic kidney diseases | | | |
| Chronic kidney disease (CKD) | 1886 (30.3) | 1948 (31.3) | 0.02 |
| Hypertensive chronic kidney disease | 1241 (19.9) | 1223 (19.7) | 0.007 |
| Chronic liver disease | | | |
| Alcoholic liver disease | 131 (2.1) | 140 (2.2) | 0.01 |
| Fibrosis and cirrhosis of liver | 246 (4.0) | 268 (4.3) | 0.02 |
| Chronic passive congestion of liver | 85 (1.4) | 97 (1.6) | 0.02 |
| Portal hypertension | 119 (1.9) | 133 (2.1) | 0.02 |
| Other specified diseases of liver | 293 (4.7) | 314 (5.0) | 0.02 |
| Cerebral infarction | 920 (14.8) | 857 (13.8) | 0.03 |
| Dementia | | | |
| Vascular dementia | 329 (5.3) | 268 (4.3) | 0.05 |
| Dementia in other diseases classified elsewhere | 545 (8.8) | 413 (6.6) | 0.08 |
| Unspecified dementia | 1196 (19.2) | 977 (15.7) | 0.09 |
| Alzheimer disease | 368 (5.9) | 298 (4.8) | 0.05 |
| Frontotemporal dementia | 23 (0.4) | 19 (0.3) | 0.01 |
| Dementia with Lewy bodies | 44 (0.7) | 34 (0.5) | 0.02 |
| Neoplasms | | | |
| Neoplasms (any) | 1790 (28.8) | 1905 (30.6) | 0.04 |
| Organ transplant | | | |
| Renal Transplantation Procedures | 46 (0.7) | 57 (0.9) | 0.02 |
| Liver Transplantation Procedures | 11 (0.2) | 21 (0.3) | 0.03 |
| Psoriasis | 82 (1.3) | 79 (1.3) | 0.004 |
| Rheumatoid arthritis | | | |
| Rheumatoid arthritis with rheumatoid factor | 40 (0.6) | 36 (0.6) | 0.008 |
| Other rheumatoid arthritis | 175 (2.8) | 166 (2.7) | 0.009 |
| Systemic lupus erythematosus (SLE) | 51 (0.8) | 47 (0.8) | 0.007 |
| Disorders involving the immune mechanism | 268 (4.3) | 308 (5.0) | 0.03 |

**Table S13** – Hazard ratios and log-rank p-values estimated for the comparison between the COVID-19 cohorts with vs. without hospitalization and with vs. without when restricting the follow-up window to the 15-180 days (thus excluding the first 14 days).

| | **Hospitalization vs. not** | | | | **Encephalopathy vs. not** | | | |
|---|---|---|---|---|---|---|---|---|
| | **1-180 days** | | **15-180 days** | | **1-180 days** | | **15-180 days** | |
| | HR (95% CI) | P-value | HR (95% CI) | P-value | HR (95% CI) | P-value | HR (95% CI) | P-value |
| Intracranial haemorrhage (any) | 3.09 (2.43-3.94) | <0.0001 | 2.56 (1.96-3.33) | <0.0001 | 4.73 (3.15-7.11) | <0.0001 | 3.15 (2.06-4.81) | <0.0001 |
| Intracranial haemorrhage (first) | 3.75 (2.49-5.64) | <0.0001 | 2.62 (1.70-4.05) | <0.0001 | 5.00 (2.33-10.70) | <0.0001 | 2.45 (1.12-5.35) | 0.037 |
| Ischaemic stroke (any) | 1.65 (1.48-1.85) | <0.0001 | 1.50 (1.32-1.70) | <0.0001 | 1.65 (1.38-1.97) | <0.0001 | 1.42 (1.15-1.75) | 0.00092 |
| Ischaemic stroke (first) | 2.82 (2.22-3.57) | <0.0001 | 2.17 (1.65-2.87) | <0.0001 | 3.39 (2.17-5.29) | <0.0001 | 1.99 (1.19-3.34) | 0.017 |
| Parkinsonism | 2.63 (1.45-4.77) | 0.0016 | 2.13 (1.07-4.24) | 0.031 | 1.64 (0.75-3.58) | 0.24 | 0.98 (0.32-2.95) | 0.6 |
| Guillain-Barre syndrome | 2.94 (1.60-5.42) | 0.00094 | 2.06 (1.15-3.68) | 0.018 | 2.27 (0.76-6.73) | 0.24 | 3.29 (1.25-8.67) | 0.16 |
| Nerve/nerve root/plexus disorders | 0.94 (0.83-1.06) | 0.29 | 0.96 (0.84-1.10) | 0.56 | 1.41 (1.07-1.87) | 0.018 | 1.44 (1.05-1.98) | 0.027 |
| Myoneural junction/muscle disease | 7.76 (5.15-11.69) | <0.0001 | 6.22 (4.11-9.42) | <0.0001 | 5.40 (3.21-9.07) | <0.0001 | 8.46 (4.04-17.70) | <0.0001 |
| Encephalitis | 3.26 (1.75-6.06) | 0.00017 | 2.54 (1.32-4.86) | 0.0094 | 9.98 (2.98-33.43) | <0.0001 | 4.78 (1.36-16.86) | 0.018 |
| Dementia | 2.28 (1.80-2.88) | <0.0001 | 1.60 (1.21-2.13) | 0.0012 | 4.25 (2.79-6.47) | <0.0001 | 4.09 (2.40-6.98) | <0.0001 |
| Mood/Anxiety/Psychotic disorder (any) | 1.23 (1.18-1.28) | <0.0001 | 1.08 (1.03-1.13) | 0.0012 | 1.73 (1.58-1.90) | <0.0001 | 1.60 (1.44-1.78) | <0.0001 |
| Mood/Anxiety/Psychotic disorder (first) | 1.55 (1.40-1.71) | <0.0001 | 1.45 (1.29-1.64) | <0.0001 | 2.28 (1.80-2.89) | <0.0001 | 2.31 (1.69-3.15) | <0.0001 |
| Mood disorder (any) | 1.21 (1.15-1.28) | <0.0001 | 1.06 (0.99-1.12) | 0.077 | 1.51 (1.35-1.70) | <0.0001 | 1.34 (1.17-1.53) | <0.0001 |
| Mood disorder (first) | 1.53 (1.33-1.75) | <0.0001 | 1.40 (1.19-1.65) | <0.0001 | 2.09 (1.55-2.80) | <0.0001 | 2.15 (1.49-3.09) | <0.0001 |
| Anxiety disorder (any) | 1.16 (1.10-1.22) | <0.0001 | 1.05 (0.99-1.11) | 0.11 | 1.64 (1.45-1.84) | <0.0001 | 1.63 (1.42-1.87) | <0.0001 |
| Anxiety disorder (first) | 1.49 (1.34-1.65) | <0.0001 | 1.38 (1.22-1.57) | <0.0001 | 1.91 (1.48-2.45) | <0.0001 | 2.25 (1.61-3.14) | <0.0001 |
| Psychotic disorder (any) | 2.22 (1.92-2.57) | <0.0001 | 2.00 (1.69-2.36) | <0.0001 | 3.84 (2.90-5.10) | <0.0001 | 2.89 (2.12-3.94) | <0.0001 |
| Psychotic disorder (first) | 2.77 (1.99-3.85) | <0.0001 | 3.26 (2.12-5.03) | <0.0001 | 5.62 (2.93-10.77) | <0.0001 | 3.44 (1.68-7.02) | 0.0013 |
| Substance misuse (any) | 1.53 (1.42-1.64) | <0.0001 | 1.32 (1.21-1.44) | <0.0001 | 1.45 (1.24-1.70) | <0.0001 | 1.49 (1.24-1.79) | <0.0001 |
| Substance misuse (first) | 1.68 (1.40-2.01) | <0.0001 | 1.39 (1.11-1.75) | 0.0042 | 2.03 (1.32-3.11) | 0.0015 | 1.79 (0.99-3.22) | 0.058 |
| Insomnia (any) | 1.08 (0.99-1.18) | 0.088 | 1.02 (0.92-1.12) | 0.76 | 1.73 (1.42-2.11) | <0.0001 | 2.11 (1.67-2.67) | <0.0001 |
| Insomnia (first) | 1.49 (1.28-1.74) | <0.0001 | 1.53 (1.29-1.83) | <0.0001 | 3.44 (2.35-5.04) | <0.0001 | 3.07 (2.03-4.63) | <0.0001 |

**Table S14** – Mean (standard deviation) of the number of visits that each cohort of patients has received during the follow-up period. Visits are counted for the matched cohorts and since the matched COVID-19 cohort varies from one comparison to the other, so does the number of visits.

| | **COVID-19 cohort mean (SD) number of visits** | **Control cohort mean (SD) number of visits** |
|---|---|---|
| Influenza | 4.05 (7.36) | 4.23 (8.09) |
| Other RTI | 3.94 (7.45) | 4.21 (8.00) |
| Skin infection | 4.13 (7.67) | 5.05 (9.24) |
| Urolithiasis | 4.49 (8.07) | 5.27 (8.51) |
| Fracture or a large bone | 3.96 (7.51) | 5.89 (9.19) |
| Pulmonary embolism | 6.14 (9.70) | 10.55 (15.25) |
